# Monoallelic *POLR3A* variants cause a Pol III-related disorder characterized by peripheral neuropathy

**DOI:** 10.1101/2025.07.16.25331559

**Authors:** Luiza L. P. Ramos, Jevin M. Parmar, Robin Wijngaard, Bianca R. Grosz, Tamas Lazar, Ligia Mateiu, Steve Vucic, Kishore R. Kumar, Dennis Yeow, Laura I. Rudaks, Lonneke de Boer, Annemarie de Vreugd, David A. Koolen, Thatjana Gardeitchik, Anita Cairns, Krishnan Iyengar, Fernando Kok, Fernanda Barbosa Figueiredo, Alzira Alves de Siqueira Carvalho, Luiz Sergio Mageste Barbosa, Rodrigo Rezende Arantes, Tyler Rehbein, Jordan E. Bontrager, Elizabeth P. Wood, Janet E. Sowden, Ivy Cuijt, Melina Ellis, Gonzalo Perez-Siles, Elyshia McNamara, Ronald van Beek, Celine B. Meijers, Ivaylo Tournev, Stephan Zuchner, Shoshana J. Wodak, Clara D.M. van Karnebeek, Nigel Laing, Liana N. Semcesen, David A. Stroud, David N. Herrmann, Velina Guergueltcheva, Marina L. Kennerson, Machteld M. Oud, Gianina Ravenscroft, Ayse Candayan, Albena Jordanova

**Affiliations:** Molecular Neurogenomics Group, VIB Center for Molecular Neurology, VIB, 2610 Antwerp, Belgium; Department of Biomedical Sciences, University of Antwerp, 2610 Antwerp, Belgium; Harry Perkins Institute of Medical Research, Centre for Medical Research, University of Western Australia, 6009 Nedlands, WA, Australia; Department of Human Genetics, Radboud university medical center, 6525 Nijmegen, The Netherlands; Donders Institute for Brain, Cognition and Behaviour, Radboud university medical center, 6525 Nijmegen, The Netherlands; United for Metabolic Diseases (UMD), The Netherlands; Northcott Neuroscience Laboratory, ANZAC Research Institute, 2139 Sydney Local Health District, NSW, Australia; School of Medical Sciences, Faculty of Medicine and Health, The University of Sydney, 2050 Camperdown, NSW, Australia; VIB-VUB Center for Structural Biology, VIB, 1050 Brussels, Belgium; Structural Biology Brussels (SBB), Department of Bioengineering, Vrije Universiteit Brussel (VUB), 1050 Brussels, Belgium; Department of Medical Genetics, University of Antwerp, 2650 Antwerp, Belgium; Brain and Nerve Research Center, Concord Clinical School, University of Sydney, 2139 NSW, Australia; Neurology Department, Concord Repatriation General Hospital, Sydney Local Health District, 2139 NSW, Australia; Neurology Department and Molecular Medicine Laboratory, Concord Repatriation General Hospital, Sydney Local Health District and NSW Health Pathology, 2139 Sydney, NSW, Australia; Translational Neurogenomics Group, Genomic and Inherited Disease Program, The Garvan Institute of Medical Research, 2010 Darlinghurst, NSW, Australia; School of Clinical Medicine, UNSW Medicine & Health, St. Vincent’s Healthcare Clinical Campus, Faculty of Medicine and Health, UNSW Sydney, 2010 Sydney, NSW, Australia; Sydney Medical School, University of Sydney, 2006 Camperdown, NSW, Australia; Clinical Genetics Unit, Royal North Shore Hospital, 2065 Sydney, NSW, Australia; Department of Pediatrics, Amalia Children’s Hospital, Radboud university medical center, 6525 Nijmegen, The Netherlands; Neurosciences Department, Queensland Children’s Hospital, 4101 Brisbane, QLD, Australia; Department of Anatomical Pathology, Royal Brisbane and Women’s Hospital. 4006 Brisbane, QLD, Australia; Department of Neurology, University of Sao Paulo School of Medicine, 05403-000 Sao Paulo, SP, Brazil; Mendelics Genomic Analysis, 02511-000 Sao Paulo, SP, Brazil; Neuromuscular Reference Center, Citadella Hospital, University of Liege, 4000 Liege, Belgium; Neuromuscular Disease Centre, Department of Neurosciences, University Center ABC, 09060-650 Santo André, SP, Brazil; Neuromuscular Disease Centre, Department of Neurology, University Hospital of the Federal University of Minas Gerais, 30130-100 Belo Horizonte, MG, Brazil; Medical Genetics Service, University Hospital of the Federal University of Minas Gerais, 30130-100 Belo Horizonte, MG, Brazil; Department of Neurology, University of Rochester, 14642 Rochester, NY, USA; Department of Neurology, Clinic of Nervous Diseases, UMBAL Aleksandrovska, Medical University-Sofia, 1431 Sofia, Bulgaria; Department of Cognitive Science and Psychology, New Bulgarian University, 1618 Sofia, Bulgaria; Dr. John T. Macdonald Foundation Department of Human Genetics, University of Miami Miller School of Medicine, 33136 Miami, FL, USA; John P. Hussman Institute for Human Genomics, University of Miami Miller School of Medicine, 33136 Miami, FL, USA; Department of Pediatrics and Human Genetics, Emma Center for Personalized Medicine, Amsterdam University Medical Center, 1105 Amsterdam, The Netherlands; Department of Biochemistry and Pharmacology, Bio21 Molecular Science and Biotechnology Institute, University of Melbourne, 3052 Parkville, VIC, Australia; Murdoch Children’s Research Institute, 3052 Melbourne, VIC, Australia; Victorian Clinical Genetics Services, 3052 Melbourne, VIC, Australia; Clinic of Neurology, University Hospital Sofiamed, 1797 Sofia, Bulgaria; Department of Neurology, Sofia University “St. Kliment Ohridski”, 1504 Sofia, Bulgaria; Molecular Medicine Laboratory, Concord Repatriation General Hospital, 2139 Concord, NSW, Australia; Molecular Medicine Center, Department of Medical Chemistry and Biochemistry, Medical University-Sofia, 1431 Sofia, Bulgaria

**Keywords:** POLR3-related disorders, Charcot-Marie-Tooth disease, CMT, RNA metabolism, peripheral nerve degeneration, transcriptomics

## Abstract

RNA polymerase III (Pol III) is a multi-subunit enzymatic complex essential for the transcription of small noncoding RNAs with structural, translational or regulatory functions. Biallelic pathogenic variants in multiple genes encoding Pol III subunits have been linked to a spectrum of neurological disorders, mainly affecting the central nervous system. To date, pathogenic monoallelic variants have been reported in only one subunit gene (*POLR3B*) in association with a spectrum of neurodevelopmental disorders, epilepsy and peripheral neuropathy.

Here, we describe a novel clinical and genetic Pol III-related entity associated with monoallelic missense variants in *POLR3A,* and presenting primarily with peripheral neuropathy. We identified eleven patients across eight unrelated families harbouring pathogenic heterozygous missense variants in the *POLR3A* gene occurring either *de novo* or segregating in an autosomal dominant fashion. Systematic clinical evaluation revealed the patients present with an early onset, progressive sensorimotor peripheral polyneuropathy with intermediate to demyelinating ranges of nerve conduction slowing and occasionally accompanied with additional neurological or non-neurological features. White matter abnormalities, characteristic for the biallelic Pol III-related disorders, were not observed in the brain magnetic resonance imaging from available individuals. Structural modelling revealed the neuropathy-associated variants cluster in regions critical for the canonical function of the polymerase, and do not overlap with known biallelic disease-causing variants. Follow up transcriptomic studies in patient-derived cells demonstrated impaired Pol III activity, including mis-regulation of individual Pol III targets and global downregulation of tRNA pools. The Pol III dysfunction was not due to impaired POLR3A expression, subcellular localization or subunit interactions.

Our findings highlight the importance of recognizing monoallelic *POLR3A* variants as a cause of peripheral neuropathy. We provide critical insights into the diversity of *POLR3A*-related allelic disorders offering a framework to understanding their underlying molecular pathomechanisms. This work highlights a central role of RNA Polymerase III dysfunction in human disease and further strengthens the link between tRNA metabolism and peripheral neurodegeneration.

## Introduction

Eukaryotic RNA Polymerase III (Pol III) is a 17-subunit enzymatic complex responsible for the transcription of small noncoding RNAs including 5S rRNA, nuclear-encoded tRNAs, and several regulatory RNAs. 1,2 Biallelic pathogenic variants in multiple genes encoding subunits of Pol III have been reported to cause heterogeneous clinical phenotypes primarily affecting the central nervous system. Recessive genetic alterations in *POLR3A*,^3–5^ *POLR3B*,^5,6^ *POLR1C*,^7^ *POLR3K*^8^ and *POLR3GL*^9^ subunits lead to hypomyelinating leukodystrophy (HLD) and often include extra-neurological features including abnormal dentition, endocrine and ocular abnormalities. Biallelic variants in *POLR3A*,^10–13^ *POLR3B*,^14^ and *POLR3GL*^15^ also cause Wiedemann-Rautenstrauch syndrome (WRS), a neonatal progeroid disorder characterized by dysmorphic facial features, intrauterine growth restriction, lipodystrophy, tremor, ataxia, global developmental delay, intellectual disability and post-natal failure to thrive.^16^ Hypomorphic *POLR3A* variants have also been reported to cause spastic ataxia and spastic paraparesis when compound heterozygous with a pathogenic *POLR3A* variant.^17–20^

Whilst biallelic variants in Pol III subunit genes are well-established as a cause of inherited neurological disease, pathogenic monoallelic variants have been reported in *POLR3B* only.^21–23^ The affected individuals present with global developmental delay, epilepsy, ataxia and progressive spasticity. Early-onset, demyelinating, sensorimotor peripheral neuropathy is frequently observed as part of the syndrome and, occasionally, can be the only clinical presentation, classified as Charcot-Marie-Tooth type 1I (CMT1I, MIM #619742). POLR3B constitutes the active site of Pol III together with POLR3A, the largest subunit in the enzymatic complex.^24^ Despite their shared involvement in forming the catalytic core, monoallelic pathogenic *POLR3A* variants have not previously been associated with a neurological phenotype.

Here, we report monoallelic *POLR3A* missense variants as a novel cause of peripheral neuropathy. We identified eleven patients across eight unrelated families, two with autosomal dominant inheritance and six with *de novo* variants, who consistently presented with early onset, sensorimotor polyneuropathy either in isolation, or with variable central nervous system (CNS) or non-neurological features. Notably, this cohort had no white matter abnormalities on brain MRI, in contrast to patients carrying biallelic pathogenic *POLR3A* variants. Structural modelling suggested that the affected residues localize to domains critical for Pol III enzymatic activity or nucleic acid interaction, while transcriptome analysis revealed dysregulation of known Pol III target genes and a global reduction of the cellular tRNA pool, consistent with impaired Pol III activity. Our findings broaden the genetic and phenotypic spectrum of *POLR3A*-related neurological disorders and have important implications for the diagnostic evaluation of patients with peripheral neuropathies.

## Materials and methods

### Patient cohort

The patient cohort was established through collaboration with the Inherited Neuropathy Consortium, diagnostic laboratories, GeneMatcher,^25^ and GENESIS GenePair tools.^26^ Written informed consent was obtained from all participants or a legal guardian according to the Declaration of Helsinki, with the study being approved by local ethics committees of the institutes involved. The participants underwent systematic and standardized physical, neurological, and electrophysiological examinations in their referring clinical centres. Whenever available, neuroradiological data was obtained.

### Genetic analyses

Genomic DNA from the probands and available family members was isolated from peripheral blood or buccal swab samples according to standard procedures. Exome sequencing (ES), genome sequencing (GS), or long-read genome sequencing (lrGS) using Oxford Nanopore Technologies was performed using established protocols. Each referring centre performed independent downstream bioinformatic analyses detailed in the Supplementary Material.

### *In silico* analyses

Protein Data Bank in Europe Knowledge Base (PDBe-KB)^27^ was consulted for the three-dimensional (3D) structure of the POLR3A protein in association with the RNA polymerase III elongation complex. PyMol Molecular Graphics System^28^ versions 2.5 and 3.1 was used for the visualization of the retrieved PDB entries (PDB: 7AE3, 7FJJ, 7D59). Residue Interaction Network Generator (RING) tool^29^ v4 was used to calculate and characterize different types of intramolecular and intermolecular residue-residue interactions in the cryo-EM structure of human Pol III complex. Relative surface accessibility scores of individual residues were calculated on PDB:7AE3 using the FreeSASA software v.2.0.324 with default parameters.^30^ Additional computational analyses, including AlphaMissense^31^ pathogenicity score comparison and assessment of amino acid biochemical properties of substituted residues, are provided in the Supplementary Material.

### Cell lines

Skin-derived fibroblasts obtained from the affected individuals A-I.1, A-II.1, A-II.2, B-II.1, E-II.1, G-II.7, and six unrelated controls were cultured using standard protocols at 37°C, in humidified air containing 5% CO_2_, with passaging when >80% confluent. Peripheral blood mononuclear cells from individuals of Family F (F-I.1, F-I.2, F-II.1) and Family H (H-I.1, H-I.2, H-II.1) were extracted using a Ficoll-Paque gradient and transformed with Epstein-Barr virus (EBV). Lymphoblast lines were maintained under humidified air at 37°C and 6% CO_2_ and passaged every five days. HEK293FT cells used for the proteomics analyses were cultured using standard protocols at 37°C in humidified air (5% CO_2_), with passages performed when 70-80% confluent.

### Immunoblotting

Cells were pelleted and lysed in RIPA lysis buffer (ThermoFisher). Protein quantification in lysates was performed using the Pierce BCA protein assay kit (ThermoFisher). Total protein extracts (30 µg) were size-separated by SDS-PAGE, transferred onto a nitrocellulose membrane and incubated with primary or secondary antibodies. The blots were developed using Pierce ECL Plus substrate (ThermoFisher) and imaged. The following antibodies were used: anti-POLR3A (Abcam #ab96328, 1:1,000 dilution), anti-GCN2 (Cell Signaling #3302, 1:1,000 dilution), anti-GCN2-phospho-T899 (Abcam #ab75836, 1:1,000 dilution), anti-eIF2a (Abcam #ab26197, 1:1,000 dilution), anti-eIF2a-phospho-Ser51 (Cell Signaling #9721, 1:1,000 dilution), anti-ATF4 (ProteinTech #10835-1-AP, 1:1,000 dilution), anti-puromycin (Merck #MABE343, 1:1,000 dilution), anti-α-tubulin (Abcam #ab7291, 1:5,000 dilution), and anti-FLAG M2 (Sigma #F3165-1MG, 1:10,000 dilution).

### Immunofluorescence assay

Fibroblasts from individual E-II.1 were cultured on glass coverslips to approximately 90% confluency. The cells were fixed using ice-cold methanol for 10 min at 4°C and blocked in 2% BSA/PBS for 20 min at room temperature. The coverslips were then incubated with the primary anti-POLR3A antibody (Abcam #ab9632, 1:250 dilution). After washing, the coverslips were incubated with a fluorescently labelled secondary goat-anti-rabbit Alexa Fluor 488 antibody (Life Technologies) at a 1:400 dilution. Next, the coverslips were mounted in Fluoromount-G containing DAPI (ITK Diagnostics). Coverslips were analysed with a Zeiss Axio Imager Z2 fluorescence microscope and the images were obtained with ZEN 3.8 software (Zeiss).

### Affinity purification coupled with mass spectrometry (AP-MS)

Detailed methods are provided in the Supplementary Material. Briefly, selected variants were introduced using site-directed mutagenesis into the cDNA of *POLR3A*, cloned into an expression vector with a C-terminal FLAG epitope, and the DNA plasmids were transiently transfected into HEK293FT cells. Cells were harvested, washed, and snap-frozen on dry ice. The cell pellets were solubilised in a digitonin-containing buffer and protein concentration was determined with the Pierce BCA Protein Assay Kit (ThermoFisher). Samples underwent affinity purification to capture FLAG-interacting proteins, as previously described^32^ with modifications described in the Supplementary Material. Eluted proteins were processed for mass spectrometry and underwent liquid chromatography (LC)-tandem mass spectrometry (MS/MS) on a Q Exactive HF-X mass spectrometer (ThermoFisher) coupled with an Ultimate 3000 RSLC nanoHPLC (Dionex Ultimate 3000), operating on data-dependent acquisition mode.

Label-free quantification (LFQ) intensities for each protein were obtained and analysed using the MaxQuant platform (version 1.6.10.43)^33^ against the UniProt human protein database (exported March 2021).^34^ Subsequent analysis was performed using Perseus (version 1.6.15.0).^35^ Each experimental sample (wild-type or variant cell line) was compared to the untransfected control independently. Proteins quantified in at least two experimental sample replicates were included, and missing values in the untransfected control were imputed based on the normal distribution. A two-sided *t*-test was performed and visualised using the scatter-plot function, with significance set to *p-*value=0.05 and log_2_ fold-change=1.

### Quantitative reverse transcription polymerase chain reaction (qRT-PCR)

RNA was extracted using the miRNeasy kit (Qiagen), followed by DNase treatment using TURBO DNA-free kit (Invitrogen). RNA concentration and purity were measured using Nanodrop spectrophotometer (ThermoFisher). cDNA was synthesized using the Superscript III reverse transcriptase (ThermoFisher) or iScript cDNA synthesis kit (Bio-Rad) according to the manufacturer’s instructions. The relative expression levels of target genes were determined by qRT-PCR with *PSMB6*, *SDHA*, and *COPS7A* housekeeping genes used for normalization. The qRT-PCR was performed using SYBR™ Green PCR master mix (Applied Biosystems), SensiMix™ SYBR® Hi-ROX Kit (Meridian Bioscience) or GoTaq® Green Master Mix (Promega). Expression quantification was performed using the qBase+ software (Biogazelle). The primers used for qRT-PCR are listed in the Supplementary Table 1.

### Transcriptomics

Detailed protocols and the analysis pipeline are given in the Supplementary Material. Briefly, small RNA enriched samples were extracted from the EBV-transformed lymphoblasts of three affected (F-I.1, F-II.1, H-II.1) and three unaffected individuals (F-II.1, H-1.1, H-1.2) from two families using miRNeasy kit (Qiagen). Two small RNAs (70 and 94 nucleotides) were synthesized and spiked into the sequencing libraries alongside the standard External RNA Controls Consortium (ERCC) mix. Libraries were prepared according to the Illumina Stranded Total RNA preparation protocol with ligation using the Ribo-Zero Plus kit with less stringent clean-up steps (1.2x). Sequencing was performed on an Element Biosciences AVITI platform.

Quality control, end trimming, mapping to human reference hg38, mapping quality assessment, gene count extraction, and expression normalization were performed following standard protocols. Gene counts were extracted in three successive analyses using *1)* all mapped reads, *2)* only primary alignments, and *3)* only uniquely mapped reads (see Supplementary Material). Differential expression analysis was done using DESeq2.^36^

### Statistical analyses

GraphPad Prism 10.4.0 was used for statistical analyses. Continuous variables were compared with a two-tailed *t*-test (paired or unpaired as appropriate) for two-group comparisons, or a one-way ANOVA followed by Tukey’s test multiple group comparisons. Significance levels are indicated as follows: ns: not significant, *:p<0.05, **:p<0.01, ***:p<0.001, ****:p<0.0001.

## Data availability

Data generated during this study are included in the manuscript and its Supplementary Material. Further information can be requested from the corresponding authors.

## Results

### Identification of *POLR3A-*associated peripheral neuropathy

Exome/genome sequencing of local cohorts of genetically unresolved patients with neuromuscular or neurometabolic disorders identified heterozygous variants in *POLR3A* in four probands by research teams in Belgium, Australia (Perth), and the Netherlands (Figure 1). Remarkably, all probands exhibited a clinical phenotype characterized by a common feature of peripheral neuropathy (Table 1, Supplementary Table 2). This clinical overlap prompted a targeted search for additional patients diagnosed with peripheral neuropathy harbouring rare deleterious variants in *POLR3A*. In this way, four additional families were identified in Australia (Sydney), Brazil, and the USA. In all cases, the heterozygous variant was a missense substitution absent in gnomAD v4, having a likely pathogenic prediction by AlphaMissense, and with segregation consistent with *de novo* or autosomal dominant inheritance. Where possible, parentage was verified in families with *de novo* variants. Additionally, the variant shared by the two unrelated probands (C-II.1 and D-II.1) was shown to occur on different chromosomal backgrounds (Supplementary Figure 1 and 2). Given the established association of *POLR3A* with recessive disorders, the intronic and untranslated regions of the gene were screened with no additional pathogenic variants being identified in any of the probands.

**Figure 1:**
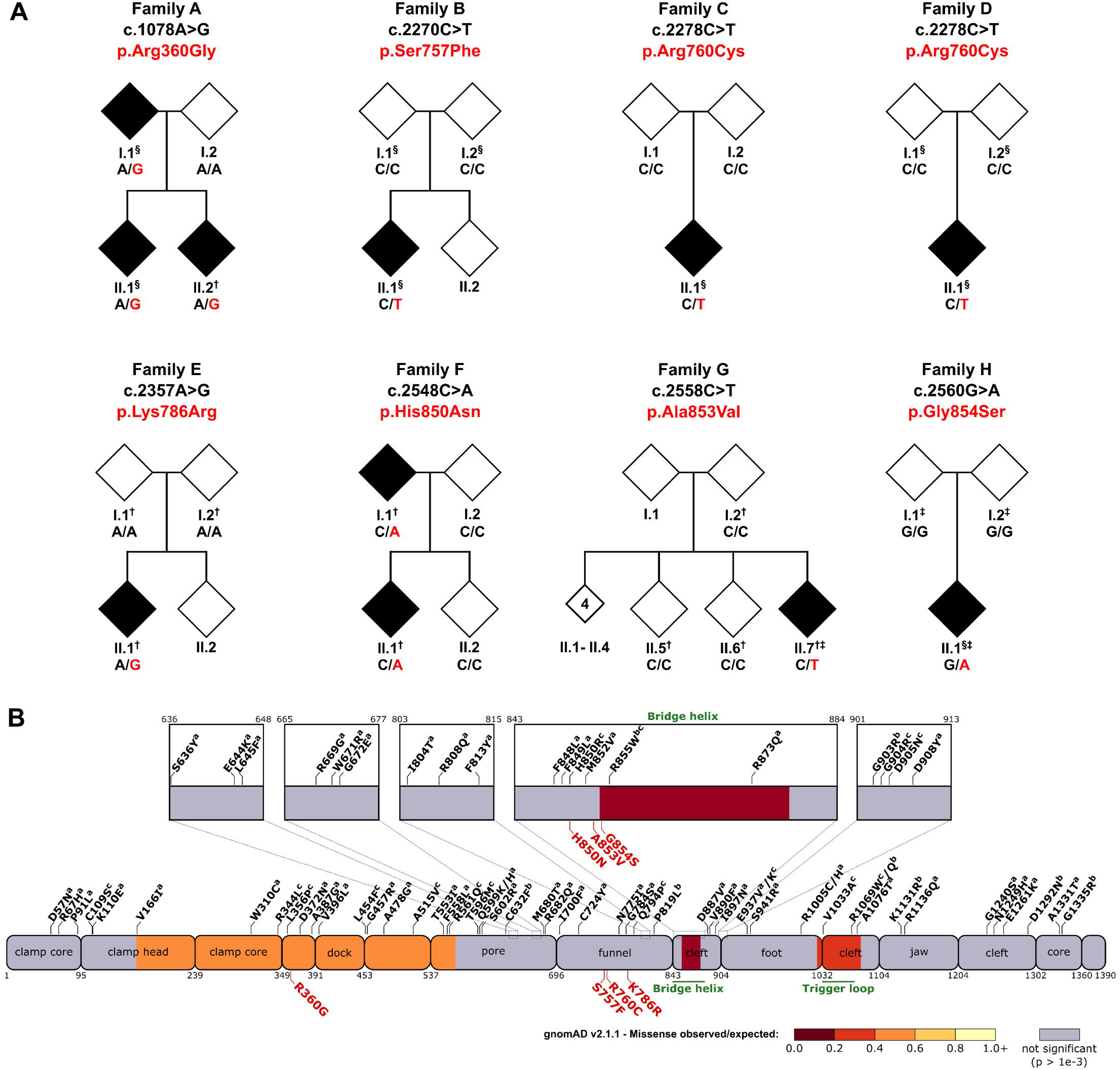
Heterozygous missense variants in *POLR3A* cause peripheral neuropathy. A: Segregation analysis in pedigrees with monoallelic *POLR3A* missense variants identified in this study. Variant identification was done using ES (^§^), GS (†), or lrGS (‡) as indicated. **B:** Schematic representation of the POLR3A protein sequence with missense substitutions linked to neurological phenotypes mapped onto annotated domains. Colour coding reflects regional missense constraint based on data from gnomAD v.2.1.1. Substitutions in black are associated with *POLR3A*-related biallelic disorders: (a) hypomyelinating leukodystrophy, (b) Wiedemann-Rautenstrauch syndrome, (c) spastic ataxia/spastic paraparesis without overt hypomyelination. Substitutions identified as disease-causing for peripheral neuropathy in this study are highlighted in red.

**Table 1.**
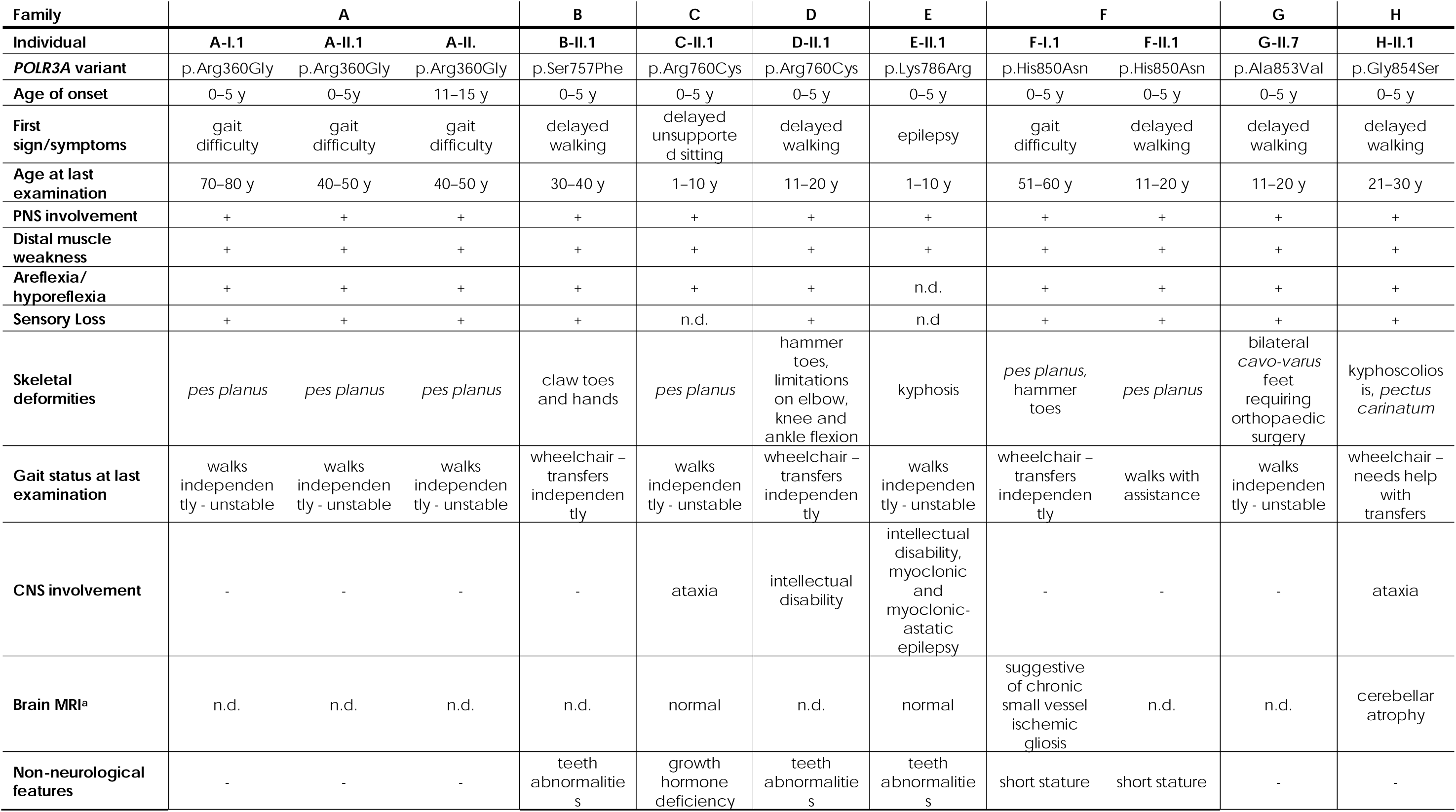
Summary of clinical characteristics of the families with pathogenic monoallelic *POLR3A* variants. + = present; - = absent; y:year or years. aage at brain MRI data is shown in parenthesis.

| Family | A |  |  | B | C | D | E | F |  | G | H |
| --- | --- | --- | --- | --- | --- | --- | --- | --- | --- | --- | --- |
| Individual | A-I.1 | A-II.1 | A-II. | B-II.1 | C-II.1 | D-II.1 | E-II.1 | F-I.1 | F-II.1 | G-II.7 | H-II.1 |
| ***POLR3A* variant** | p.Arg360Gly | p.Arg360Gly | p.Arg360Gly | p.Ser757Phe | p.Arg760Cys | p.Arg760Cys | p.Lys786Arg | p.His850Asn | p.His850Asn | p.Ala853Val | p.Gly854Ser |
| **Age of onset** | 0–5 y | 0–5y | 11–15 y | 0–5 y | 0–5 y | 0–5 y | 0–5 y | 0–5 y | 0–5 y | 0–5 y | 0–5 y |
| **First sign/symptoms** | gait difficulty | gait difficulty | gait difficulty | delayed walking | delayed unsupported sitting | delayed walking | epilepsy | gait difficulty | delayed walking | delayed walking | delayed walking |
| **Age at last examination** | 70–80 y | 40–50 y | 40–50 y | 30–40 y | 1–10 y | 11–20 y | 1–10 y | 51–60 y | 11–20 y | 11–20 y | 21–30 y |
| **PNS involvement** | + | + | + | + | + | + | + | + | + | + | + |
| **Distal muscle weakness** | + | + | + | + | + | + | + | + | + | + | + |
| **Areflexia/hyporeflexia** | + | + | + | + | + | + | n.d. | + | + | + | + |
| **Sensory Loss** | + | + | + | + | n.d. | + | n.d | + | + | + | + |
| **Skeletal deformities** | *pes planus* | *pes planus* | *pes planus* | claw toes and hands | *pes planus* | hammer toes, limitations on elbow, knee and ankle flexion | kyphosis | *pes planus*, hammer toes | *pes planus* | bilateral *cavo-varus* feet requiring orthopaedic surgery | kyphoscoliosis, *pectus carinatum* |
| **Gait status at last examination** | walks independently - unstable | walks independently - unstable | walks independently - unstable | wheelchair – transfers independently | walks independently - unstable | wheelchair – transfers independently | walks independently - unstable | wheelchair – transfers independently | walks with assistance | walks independently - unstable | wheelchair – needs help with transfers |
| **CNS involvement** | - | - | - | - | ataxia | intellectual disability | intellectual disability, myoclonic and myoclonic-astatic epilepsy | - | - | - | ataxia |
| **Brain MRI<sup>a</sup>** | n.d. | n.d. | n.d. | n.d. | normal | n.d. | normal | suggestive of chronic small vessel ischemic gliosis | n.d. | n.d. | cerebellar atrophy |
| **Non-neurological features** | - | - | - | teeth abnormalities | growth hormone deficiency | teeth abnormalities | teeth abnormalities | short stature | short stature | - | - |

All affected individuals underwent deep clinical characterization summarized in Table 1 and detailed in Supplementary Material. The cohort included nine males and two females, with ages at the last examination ranging from 7 to 78 years. Gross motor impairment was the initial and predominant symptom in most patients, typically presenting as delayed independent walking and/or gait difficulties, with a mean age of onset at 3.15 ±3.4 years. Some individuals exhibited delays in earlier motor milestones, such as crawling and unsupported sitting.

At their latest examination, symmetrical muscle weakness was present in the lower limbs of all patients and in the upper limbs of 90% of patients for whom this data was available (Supplementary Table 2). Deep tendon reflexes were absent in the lower limbs and/or distal upper limbs of all individuals assessed. Distally attenuated sensory loss, involving superficial and deep sensation was uniformly reported in all the patients tested. Motor and sensory deficits were usually more pronounced distally. Individuals D-II.1, G-II.7 and H-II.1 were underweight, a potential result of generalized muscle atrophy. Significant skeletal deformities were reported in 45% of the patients and included claw/hammer toes, *cavo-varus* feet, kyphoscoliosis, and *pectus carinatum*. Six patients had *pes planus*.

Although the cohort displayed consistent early disease onset, the effects on motor performance and disease progression varied significantly. While most patients remained ambulatory, either walking independently or with assistance, four required a wheelchair at their most recent evaluation. The disease progression variability was further exemplified in patient A-I.1, who continued to walk independently after 65 years of disease progression. In contrast, patient H-II.1 had significant impairment of both gross and fine motor function, as well as proprioception in all extremities, became wheelchair-dependent during adolescence and required assistance with transfers in his twenties.

Nerve conduction studies (NCS) were performed at ages 6 to 78 years revealing sensorimotor peripheral polyneuropathy with intermediate to demyelinating range of conduction slowing. The mean motor NCV (MNCV) in the median nerve was 28.6 m/s (range: 9-40 m/s) among tested patients (Table 2). Sensory nerve action potentials (SNAP) were undetectable in both ulnar and sural nerves in all tested patients, in accordance with the clinically observed deficits of all sensory modalities.

**Table 2.**
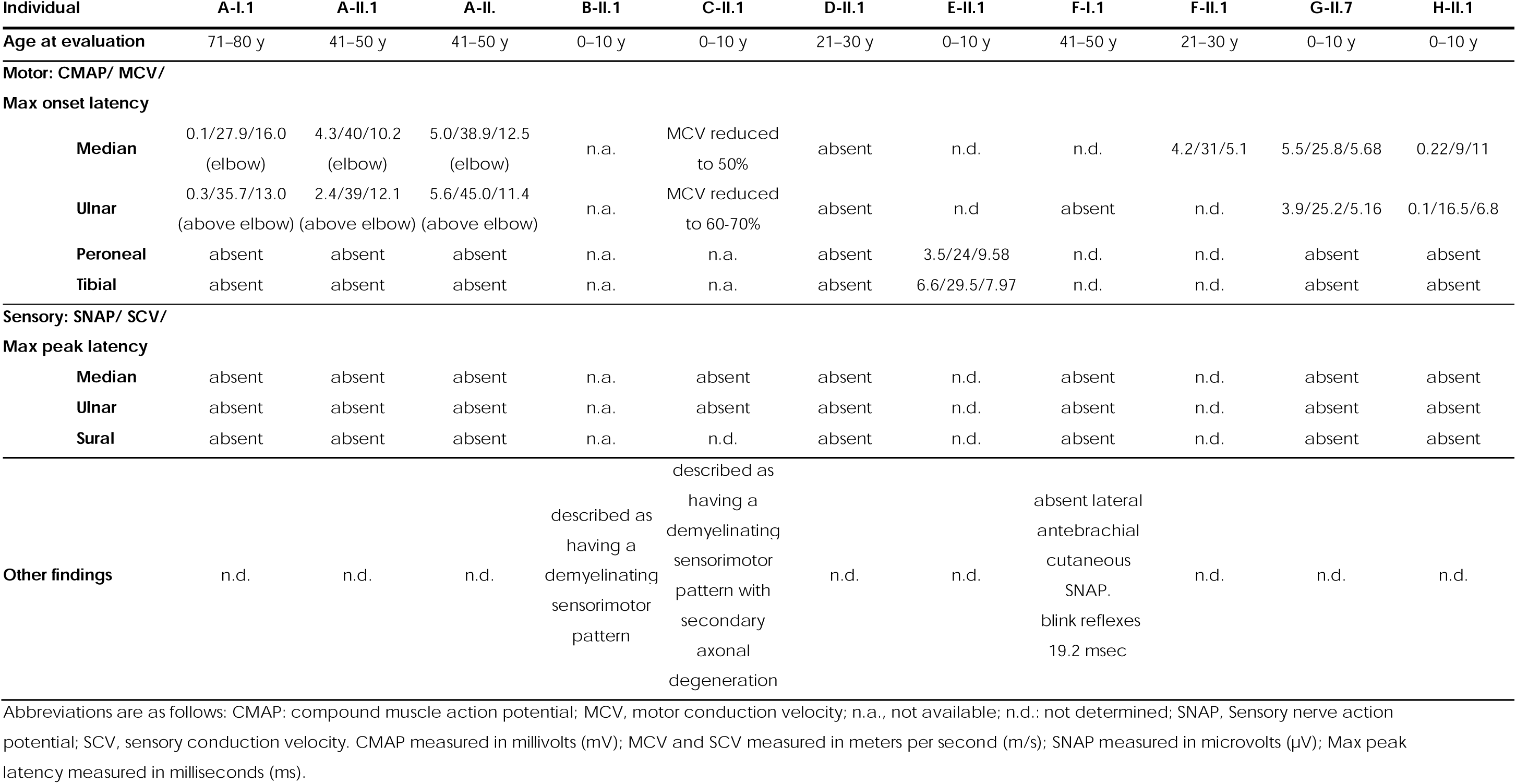
Findings from the nerve conduction studies of the families with pathogenic monoallelic *POLR3A* variants.

In addition to peripheral neuropathy, four patients demonstrated CNS involvement. Patients C-II.1 and H-II.1 displayed cerebellar ataxia, as well as dysarthria and dysphagia. Two patients (D-II.1 and E-II.1) exhibited intellectual disability. Patient E-II.1 presented with seizures as the initial symptom during infancy, which subsequently progressed to myoclonic and myoclonic-astatic epilepsy and global developmental delay.

Brain MRI data were available for four affected individuals, including three with CNS involvement (C-II.1, E-II.1, H-II.1), and one without CNS features imaged more than 50 years after disease onset (F-I.1). Notably, the neuroimaging data did not show evidence of hypomyelinating leukodystrophy or other white matter abnormalities in any of these individuals. Cerebellar atrophy was observed in one patient presenting clinical signs of ataxia (H-II.1).

Non-neurological features were observed in five affected individuals, including short stature, growth hormone deficiency, and mild teeth abnormalities (i.e. enamel defects). Interestingly, patients C-II.1 and D-II.1 who harboured the same *POLR3A* variant (p.Arg760Cys), demonstrated a similar degree of motor impairment relative to their age; however, they exhibited distinct extra-peripheral nervous system (PNS) manifestations, with the former presenting with growth hormone deficiency and the latter with intellectual disability.

### Neuropathy-associated POLR3A missense variants impair Pol III architecture and function

POLR3A, together with POLR3B, forms the active centre of Pol III where transcription takes place. Within this catalytic core, phospho-diester bonds between adjacent ribonucleotides are formed using a single-strand DNA template, while the newly formed DNA-RNA hybrid is translocated away from the transcription bubble to create space for the next ribonucleotides to arrive.^37^ As such, POLR3A needs to interact with both nucleic acids and the surrounding Pol III subunits. We performed extensive computational and experimental studies to evaluate the impact of the identified missense variants on Pol III complex architecture and function.

Modelling the spatial location of the affected residues in the cryo-EM structure of Pol III (PDB:7AE3) showed that they reside in the core of the complex, around the active site cleft (Figure 2A). These findings were corroborated by relative surface accessibility (RSA) calculations allowing estimation of the exposure of affected residues to solvent at the outer surface of the protein complex. The analysis revealed that although all affected residues are relatively solvent-accessible in the isolated POLR3A protein, their RSA scores decrease substantially when modelled within the full Pol III complex, and in the presence of nucleic acids, suggesting that these residues are buried within the assembled complex (Supplementary Figure 3A).

**Figure 2:**
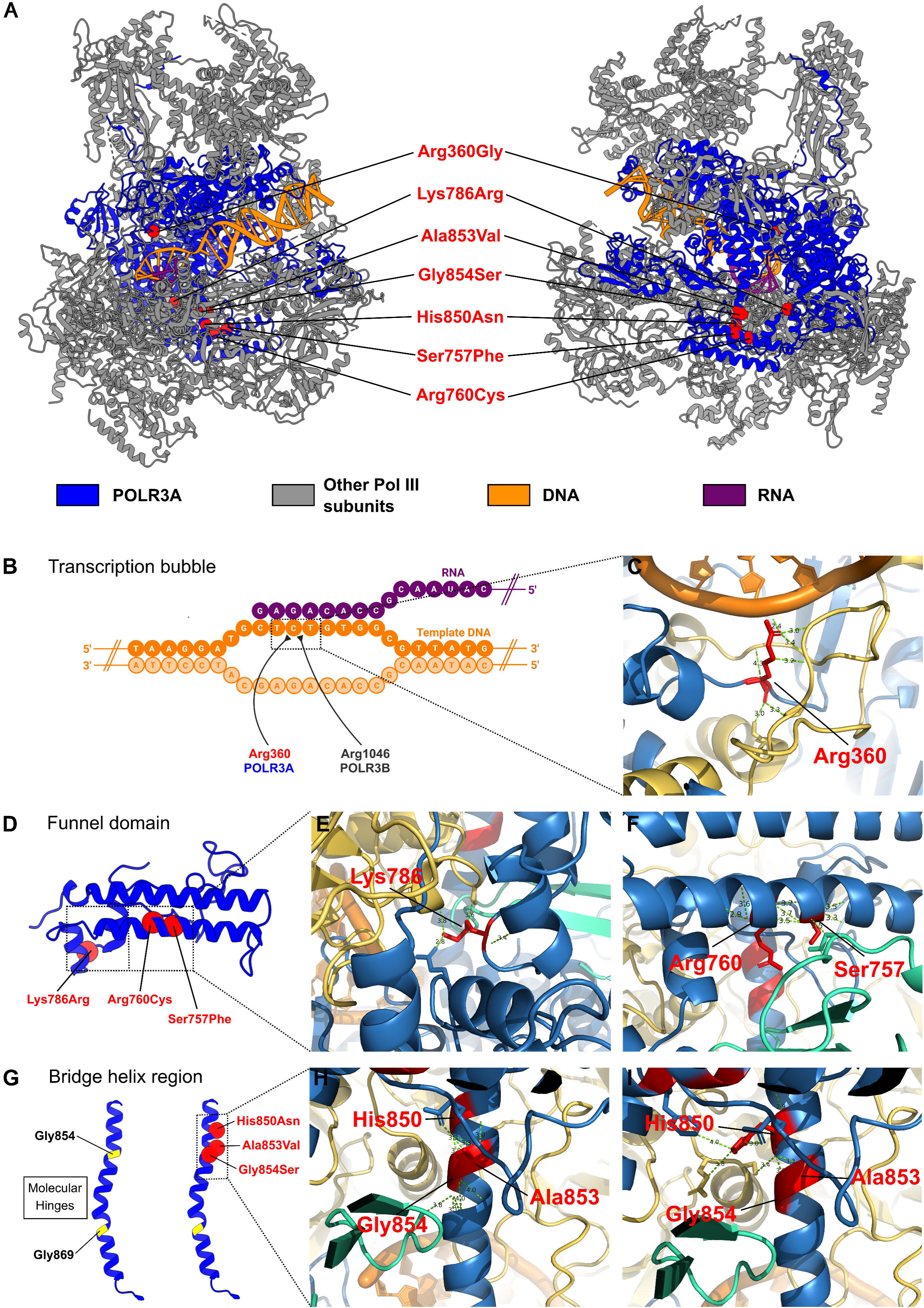
Neuropathy-associated POLR3A residues are located in regions critical for Pol III function and make stabilizing molecular interactions. A: The positions of the neuropathy-associated POLR3A residues from two viewing angles are shown on the ribbon representation of the 3D cryo-EM structure of the human Pol III complex (PDB: 7AE3). **B:** Schematic representation of the transcription bubble highlighting neuropathy-associated residues in POLR3A and POLR3B that contact the same nucleotide in the template DNA. **C, E-F, H-I:** Intramolecular and intermolecular interactions of residues likely altered by neuropathy-associated substitutions, indicated by dashed lines. In these panels, POLR3A, POLR3B, POLR3K, and neuropathy-associated residues are coloured blue, yellow, green, and red, respectively. **D:** Ribbon representation of the POLR3A funnel domain with neuropathy-associated POLR3A substitutions indicated in red. **G:** Ribbon representation of the POLR3A bridge helix (BH) region. Left: the BH with conserved glycine residues acting as molecular hinges (yellow). Right: the BH showing neuropathy-associated POLR3A substitutions in red.

Three substituted residues (p.His850, p.Ala853, p.Gly854) lie within the bridge helix region and interact with each other, while two residues (p.Ser757, p.Arg760) are located ∼16Å further from this cluster and are also involved in residue-residue interactions among themselves (Figure 2, Supplementary Table 3). The bridge helix region is a key component of the catalytic cleft and coordinates substrate movement during transcription by the kinking of two molecular hinges that are exclusively made of glycine residues.^38^ Strikingly, the neuropathy-associated p.Gly854 residue represents one of these hinges and is substituted with the bulkier serine residue in Family H (Figure 1A). Residue p.Lys786 located within the funnel domain, a narrowing channel involved in transcription termination,^39^ is predicted to make interactions within POLR3A as well as associated subunits. Finally, p.Arg360 is located within a linker region connecting the clamp core and dock domains which contributes to stabilizing DNA-RNA hybrid formation during transcription and is required for the overall stability and processivity of the elongation complex.^37,39^

In addition to *in silico* characterization, we tested the functional impact of identified substitutions in cellular systems. Prior studies in patient-derived fibroblasts^3,4^ and HeLa^40^ models of *POLR3A*-HLD have shown that certain pathogenic biallelic variants reduce POLR3A protein abundance. To investigate if this mechanism also applies to the neuropathy-associated variants in our cohort, we performed immunoblotting analysis on patient-derived cells from five families (A, B, E, F, G, H). POLR3A abundance was comparable to that of unrelated controls or unaffected family members (Figure 3A). Furthermore, immunocytochemistry of fibroblasts derived from patient E-II.1 revealed a similar subcellular POLR3A distribution to a control line, with predominant nuclear localization and some cytoplasmic staining (Supplementary Figure 4).

**Figure 3:**
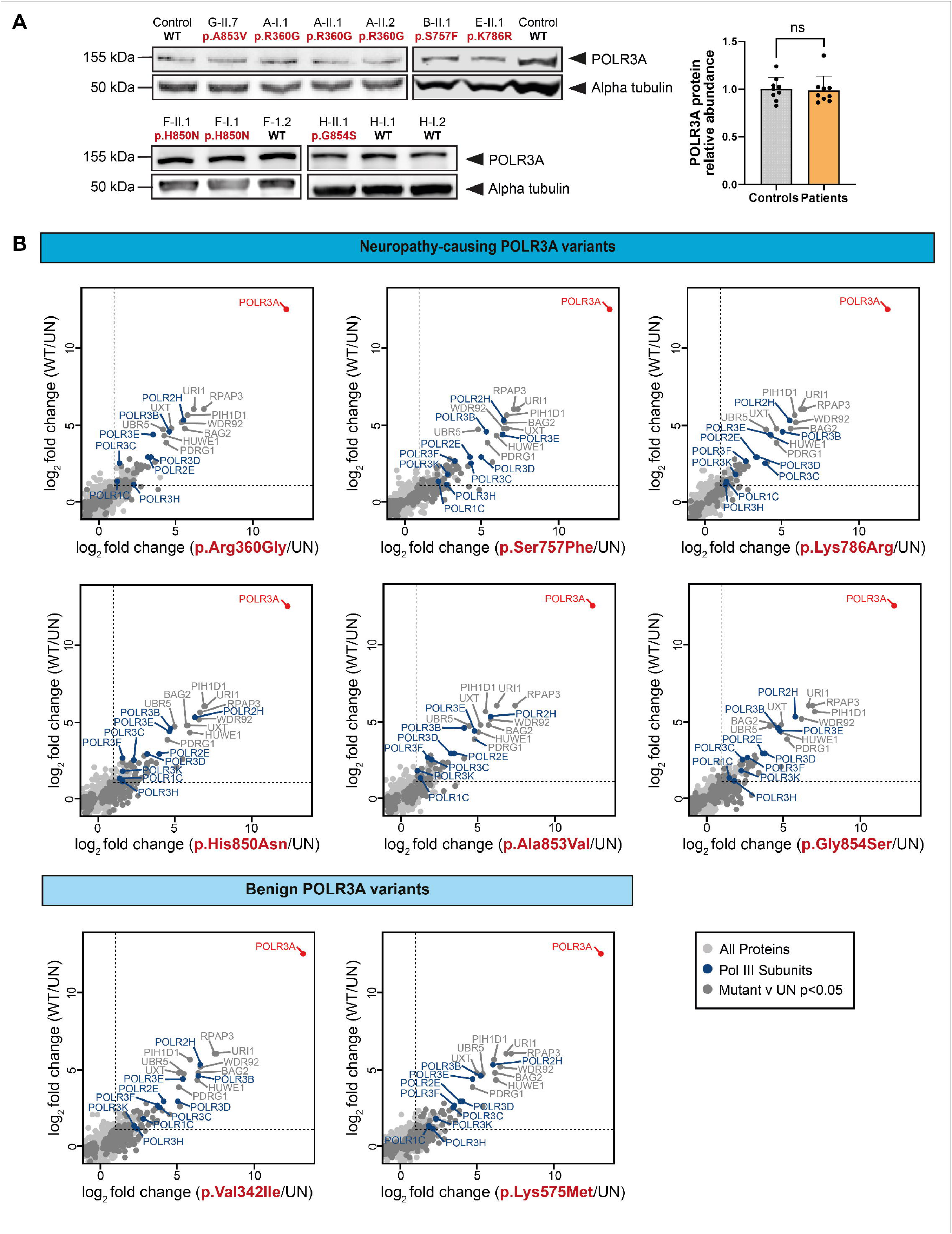
The neuropathy-associated POLR3A substitutions do not change POLR3A abundance or Pol III complex assembly. A: Western blot analysis of POLR3A protein levels in fibroblasts or lymphoblasts from patients in families A, B, E, F, G and H compared to unrelated controls or unaffected family members (left). Alpha-tubulin was used as a loading control. Quantification (right panel) shows no significant difference in POLR3A protein levels between nine patient and nine healthy control samples (unpaired *t*-test; ns = not significant). **B**: AP-MS-based interactomics profiling of FLAG-tagged POLR3A isoforms expressed in HEK293FT cells. Correlation plots show log _2_fold-change in protein abundance following enrichment by each patient isoform or benign isoform of POLR3A (x-axis) compared to wild-type (y-axis). Each condition was independently compared against untransfected (UN) controls. POLR3A is highlighted in red; other Pol III subunits are shown in dark blue. Proteins significantly enriched (mutant vs UN; *t*-test p<0.05) are indicated in dark grey. Dashed lines indicate a log_2_fold-change of 1.

To investigate the impact of the identified amino acid substitutions on the POLR3A protein-protein interactions, we performed interactome profiling for each of the neuropathy-associated alleles, except for p.Arg760Cys. For controls, we used the wild-type protein as well as two POLR3A isoforms carrying benign substitutions (p.Val342Ile or p.Lys575Met). HEK293FT cells were transiently transfected with FLAG-tagged constructs expressing either wild-type or mutant POLR3A, and interacting proteins were identified by affinity purification followed by mass spectrometry (AP-MS). Overall, ∼1,800-2,000 unique proteins were detected for each condition, including 12 of 17 Pol III subunits. As expected, POLR3A was the most abundantly precipitated protein in transfected samples relative to untransfected controls (Supplementary Figure 5A). Overall, mutant POLR3A showed strong interactions with other Pol III subunits, however these were not significantly different than interactions made by wild-type or benign POLR3A isoforms, suggesting preserved complex assembly (Figure 3B).

Although the interactomics studies suggested the overall architecture of Pol III remained intact, we questioned whether its fundamental biological function in transcription would be impaired by the neuropathy-associated *POLR3A* substitutions. Using patient-derived fibroblasts or lymphoblasts, we performed qRT-PCR analysis of four Pol III targets (5S rRNA, 7SL RNA, 7SK RNA, tRNA-Ala-TGC), each governed by a different type of promoter^41^ and reported to be dysregulated in *POLR3A*-HLD or *POLR3A*-WRS patients. BC200 RNA, a primate-specific transcript predominantly expressed in brain tissue, was also quantified due to its putative role in the hypomyelination in HLD cellular models.^40^ qRT-PCR analyses revealed differential dysregulation of the Pol III targets across families depending on the substitution location. Patients with missense variants within the funnel domain (families B and E) displayed upregulation of 7SL RNA, 5S rRNA and tRNA-Ala-TGC. In contrast, they showed no change in the BC200 RNA levels, while this transcript was consistently downregulated in all remaining patients. The patients with the variant affecting the DNA binding residue (Family A) showed a hybrid profile, characterized by elevated 7SL RNA and 5S rRNA and reduced BC200 RNA expression (Figure 4A).

**Figure 4:**
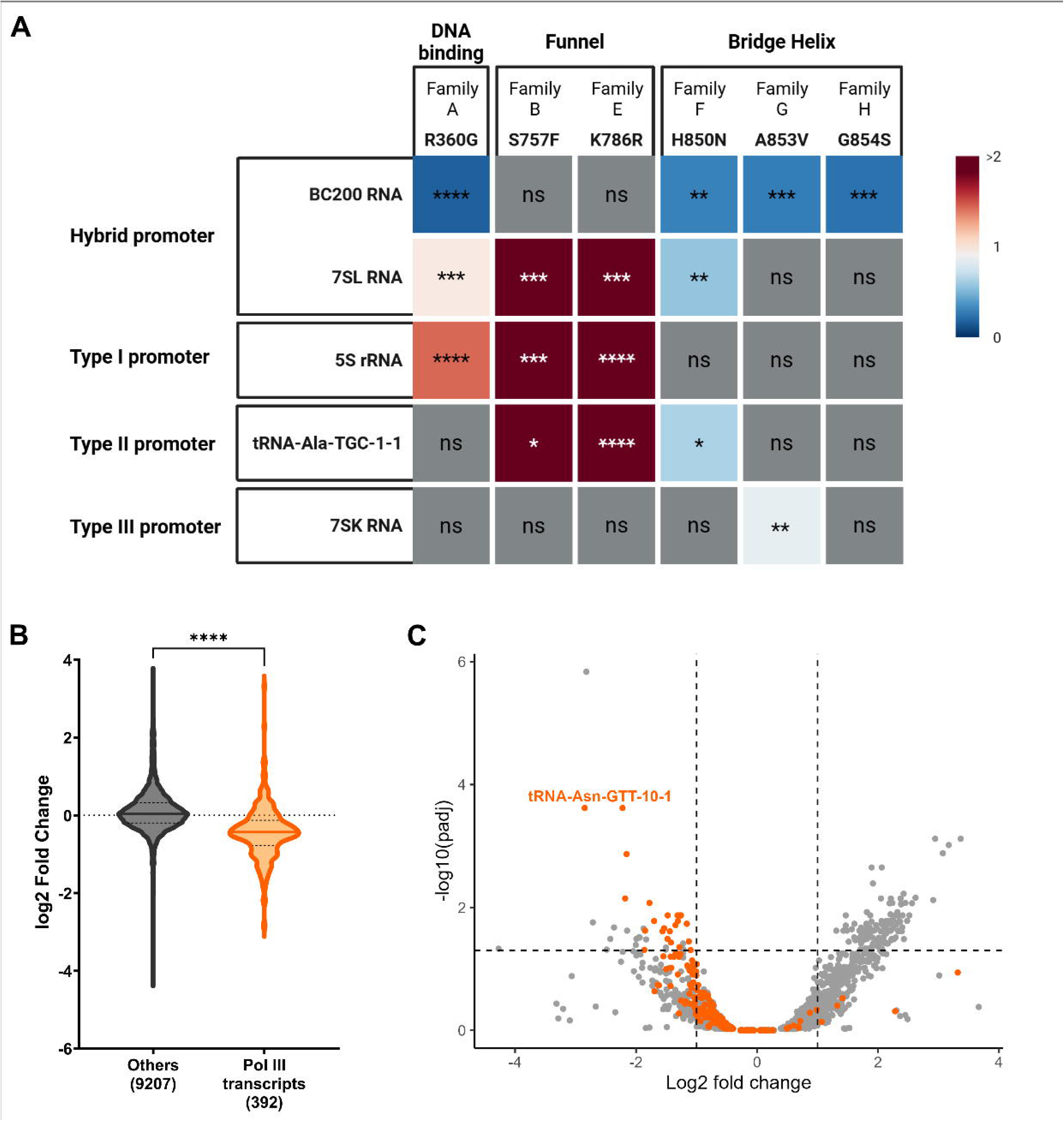
Neuropathy-associated *POLR3A* variants impair Pol III transcription activity. A: Graphical summary of qRT-PCR expression analyses for selected Pol III targets across the neuropathy cohort. Significantly downregulated targets relative to controls are shown in shades of blue; significantly upregulated targets are shown in shades of red. Darker colours indicate higher fold-change. Statistical analysis was performed using unpaired *t*-tests with Welch’s correction. **B:** Violin plot comparing the distribution of log_2_-fold change of transcripts synthesized by other major polymerases (n=9207) and transcripts synthesized by Pol III (n=392) in three patients (F-I.1, F-II.1, H-II.1) relative to three unaffected family members (F-I.2, H-I.1, H-I.2). Pol III transcripts show a significant overall downregulation compared to other major RNA polymerases (Mean_Others_=0.1005, Mean_Pol III transcripts_=-0.4436, standard error of the mean=0.02823, unpaired *t*-test with Welch’s correction). **C:** Volcano plot indicating differential expression of transcripts captured in the small RNA sequencing dataset comparing pooled expression in affected individuals (F-I.1, F-II.1, H-II.1) relative to unaffected family members (F-I.2, H-I.1, H-I.2). Each dot represents a distinct RNA transcript, with tRNAs highlighted in orange. Vertical dashed lines indicate a log_2_-fold change threshold of ±1 for significantly up-and downregulated transcripts. The horizontal dashed line marks an adjusted p-value cutoff of 0.05. One significantly downregulated transcript (tRNA-Asn-GTT-10-1) that is labelled on the plot was orthogonally validated in families F and H and investigated for differential expression in families A, B, E, and G, as well. *:p<0.05, **:p<0.01, ***:p<0.001, ****:p<0.0001, ns: non-significant.

We then conducted an unbiased small RNA transcriptome analysis in EBV-transformed lymphoblasts from three affected (F-I.1, F-II.1, H-II.1) and three unaffected individuals (F-I.2, H-I.1, H-I.2) in families F and H. An average of 89.1M reads per sample was obtained, with 87.7% meeting the Q30 quality threshold. The dataset included 9,599 distinct transcripts with >20 reads in at least three individuals. Among these, 392 were transcribed by Pol III, including 373 precursor tRNAs representing 60% of the entries listed in the tRNA database GtRNAdb.^42^ Compared to transcripts from other major RNA polymerases, Pol III transcripts showed a significant decrease in patients relative to unaffected family members, indicating a global downregulation for the Pol III transcriptome (Figure 4B). Importantly, this decrease was mainly due to tRNA depletion, with 23 tRNAs from multiple tRNA isoacceptor families significantly downregulated (adjusted p-value<0.05; Figure 4C). One significantly downregulated transcript (tRNA-Asn-GTT-10-1) was orthogonally validated in families F and H, and additionally investigated for differential expression in families A, B, E, and G using qRT-PCR. Similar to the findings in the targeted analysis, there was upregulation of tRNA-Asn-GTT-10-1 in patients with funnel domain substitutions (B-II.1 and E-II.1), while the individuals with bridge helix substitutions (F-I.1, F-II.1, G-II.7, and H-II.1) showed significant downregulation. In contrast, the patients with the DNA-binding residue substitution (A-I.1, A-II.1, A-II.2) showed no significant change in expression (Supplementary Figure 6). Taken together, these findings reinforce the variant specific trend of Pol III transcript dysregulation.

As tRNA deficiency can trigger the integrated stress response (ISR) pathway^43^ and ISR activation has been observed in the brain tissue of a *POLR3A*-HLD mouse model,^44^ we investigated key ISR markers in EBV-transformed lymphoblasts from Family F and H. Immunoblotting showed no upregulation of phospho-GCN2, phospho-eIF2a, or ATF4 in the patients relative to unaffected family members. Similarly, puromycin treatment did not reveal decreased global translation (Supplementary Figure 7).

## Discussion

This study identifies heterozygous missense variants in the *POLR3A* gene as a novel cause of early onset sensorimotor polyneuropathy. Through clinical, genetic, computational, and functional analyses we show that variants co-segregate with disease in dominant or *de novo* patterns and result in a phenotype distinct from classical *POLR3A*-related biallelic disorders.

Overall, the patients exhibited a clinically and electrophysiologically homogeneous core neuropathy phenotype, characterized by an early childhood onset, progressive sensorimotor polyneuropathy with delayed motor milestones, severe distal limb impairment, and intermediate to demyelinating range conduction slowing along peripheral nerves, consistent with a diagnosis of Charcot-Marie-Tooth neuropathy. The disease progression was generally severe, especially in the patients with *de novo* variants. All affected individuals experienced significantly impaired mobility, with approximately 40% requiring a wheelchair for ambulation within two decades of disease onset. There was inter-familial variability in the motor impairment and the rate of disease progression, leading to various degrees of muscle atrophy, motor disability, and skeletal deformities. In addition, a subset of patients exhibited features beyond the PNS to include CNS and extra-neurological involvement.

Prominent clinical CNS findings were observed in four patients including cerebellar ataxia, tremor, and intellectual disability. Brain MRI data available for these individuals showed no evidence for hypomyelinating leukodystrophy or other white matter abnormalities. Notably, patient E-II.1 (harbouring the p.Lys786Arg variant) diverged from the typical neuropathy-predominant presentation, exhibiting myoclonic-atonic seizures during infancy followed by global neurodevelopmental delay and intellectual disability. Peripheral neuropathy was not initially reported; however, the child had delayed walking and systematic neurological and electrophysiological examinations during this study revealed the presence of demyelinating sensorimotor neuropathy. Epilepsy-related gene panel testing did not identify any recurrent pathogenic variants explaining the seizures. Further cases are needed to determine if epileptic encephalopathy reflects a broader manifestation of *POLR3A*-related monoallelic disease or a coincidental co-morbidity.

Mild systemic features were observed in five patients, including teeth enamel defects or misplaced teeth, and short stature, which could be missed during routine neurological evaluations (Table 1, Supplementary Table 2). Patient C-II.1 received growth hormone treatment during childhood for delayed bone age and short stature, showing a positive response. When present alongside early-onset peripheral neuropathy, these non-neurological signs may serve as clinical clues for *POLR3A*-related neuropathy.

The clinical features observed in our *POLR3A* cohort closely resemble those reported in monoallelic *POLR3B*-related disease.^21^ To date, 23 individuals with heterozygous *POLR3B* missense variants have been described, typically presenting with peripheral neuropathy as a diagnostic hallmark. Similar extra-PNS features including ataxia, developmental delay, and endocrine abnormalities have been variably observed in both cohorts (Supplementary Table 4). As in patient E-II.1, myoclonic epilepsy has occasionally appeared as a primary symptom, with subclinical peripheral neuropathy detected only through electrophysiology.^23^ These clinical similarities support a shared clinical spectrum associated with monoallelic missense variants in both catalytic subunits of Pol III, centred around peripheral neuropathy.

Overall, there are notable differences in the clinical and radiological features of the monoallelic *POLR3A* and *POLR3B* clinical entities compared to the biallelic Pol III-related disorders. While biallelic variants commonly cause hypomyelinating leukodystrophy with typical white matter abnormalities on brain MRI, such findings were absent in all four individuals of the peripheral neuropathy cohort with available neuroimaging data, suggesting that central hypomyelination is not a core feature of the monoallelic phenotype. Similarly, none of our patients showed hallmarks of WRS, such as lipodystrophy and characteristic craniofacial dysmorphisms (Supplementary Table 4). Despite peripheral nerve involvement not being reported in patients with biallelic Pol III-related disorders,^45^ or their heterozygous carrier parents (Dr. L.S.M. Barbosa, Dr. R.R. Arantes, personal communication), sensorimotor neuropathy is the consistent feature in monoallelic Pol III-related disorders. Nevertheless, some CMT patients with extra-PNS features exhibit additional signs overlapping with recessive Pol III disorders including cerebellar ataxia, growth hormone deficiency, and enamel defects. Overall, the differential clinical and radiological hallmarks of the monoallelic vs biallelic Pol III disorders could provide valuable guidance for the variant interpretation and the molecular diagnosis of the patients.

The majority of monoallelic variants we identified cluster within the funnel domain and the bridge helix region of POLR3A.^39^ Although nine missense variants in the funnel domain and six in the bridge helix have been reported in individuals with biallelic *POLR3A*-related disorders (Figure 1B), the specific substitutions differ between the monoallelic and biallelic disease contexts. For instance, residue p.His850 is implicated in both settings: monoallelic p.His850Asn causes peripheral neuropathy; whereas p.His850Arg in compound heterozygosity with the c.1909+22G>A variant was identified in a patient presenting with ataxia, dental abnormalities and T2-hyperintense signal at the level of the posterior limb of the internal capsule.^17^ A qualitative comparison of amino acid changes suggests that neuropathy-associated substitutions are generally more structurally disruptive than those linked to *POLR3A*-HLD/WRS (Supplementary Figure 2C-D) and can cause a clinical phenotype on their own. In contrast, the substitutions associated with biallelic Pol III-related disorders are less damaging and can exert a clinical phenotype only in combination with a second, loss-of-function or hypomorphic allele. Additionally, certain residues may be intrinsically more vulnerable to change, such as p.Gly854 located at a critical hinge point in the bridge helix associated exclusively with the monoallelic phenotype. Thus, our observations suggest that monoallelic and biallelic *POLR3A*-related disorders involve non-overlapping amino acid changes, and that the phenotypic outcome may depend on the specific substitution surpassing a critical threshold of functional impairment.

We systematically characterized monoallelic *POLR3A* variants to assess their impact on Pol III function. Remarkably, *in silico* analyses revealed that all affected residues are located in the relative vicinity to the core of the polymerase complex, adjacent to the active centre, and in regions essential for transcriptional activity. For example, the p.Arg360 residue (Family A) contacts a DNA nucleotide in the transcription bubble involved in DNA-RNA hybrid stabilization.^37,39^ Intriguingly, this parallels the role of p.Arg1046 in POLR3B, a residue recurrently substituted in monoallelic *POLR3B*-related peripheral neuropathy,^21,46^ highlighting a structure-function correlation common to the *POLR3A*-and *POLR3B*-related monoallelic disorders.^47^ Additionally, a cluster of neuropathy-associated substitutions (p.Ser757Phe, p.Arg760Cys, p.Lys786Arg) localizing to the funnel domain, interact with each other, and also contribute to inter-subunit stabilization (Figure 2, Supplementary Table 3). In *Schizosaccharomyces pombe*, the p.Lys780Arg substitution, orthologous to human p.Lys786Arg, causes increased transcription termination, likely due to the enhanced interaction between the substituted arginine and a glutamate residue within the POLR3K subunit.^48^ According to a kinetic coupling model, this change would impair Pol III processivity and reduce the elongation rate, corroborating the pathogenic nature of this variant.^39,48^

Three neuropathy-associated substitutions (p.His850Asn, p.Ala853Val, p.Gly854Ser) affect neighbouring residues located in the bridge helix, a dynamic structural element critical for coordinating the conformational changes in the transcription bubble through kinking of two molecular hinges.^38^ These molecular hinges are conserved across all bacterial, archaeal and eukaryotic RNA polymerases,^38^ and are invariably made of glycine residues (Supplementary Figure 1A). Remarkably, p.Gly854 substituted in H-II.1 represents one of these molecular hinges. Due to its small size, glycine confers flexibility to the bridge helix by destabilizing its structure, making it well-suited to support the repeated bending and straightening motions required during transcription elongation.^49^ Its substitution impairs this dynamic motion, thereby disrupting efficient Pol III catalysis. Furthermore, systematic mutagenesis of the bridge helix residues in *Methanocaldococcus jannaschii* RNA polymerase showed that most amino acid substitutions, including orthologous residues identified in our cohort (p.His806Asn, p.Gly810Ser), result in reduced polymerase activity.^38^ This highlights the functional importance in maintaining the bridge helix, and supporting the deleterious impact of the identified variants.

In addition to computational analyses, direct evidence for the damaging effect of the identified variants was obtained by evaluating the Pol III transcriptional profile in patient-derived cells. Several prior studies reported differentially expressed genes (DEGs) in patients with biallelic pathogenic variants in different Pol III subunits.^8,40,50–52^ The experimental design and identity of the dysregulated Pol III targets differ among the studies, making it difficult to establish genotype-phenotype correlations. In this study, we systematically quantified the endogenous expression of five Pol III target transcripts, each corresponding to a distinct promotor type recognized by the polymerase complex, across multiple patients with monoallelic *POLR3A* variants. Although the expression pattern of the selected targets varied among analysed patients, a consistent trend emerged in transcript expression levels that correlated with the topology of the affected residues. Patients carrying variants in the funnel domain (Family B and E) showed consistent upregulation of 7SL RNA, 5S rRNA, and tRNA-Ala-TGC, but not of BC200 RNA and 7SK RNA. In contrast, patients carrying variants outside of this region had downregulation of BC200 transcript levels. This primate-specific neural transcript is reported to be downregulated in oligodendroglial cellular models and fibroblasts from *POLR3A*-HLD patients carrying the compound heterozygous p.Met852Val in combination with a null allele.^40^ Interestingly, this substitution resides in the bridge helix region and is adjacent to the three monoallelic neuropathy-associated variants. Importantly, Choquet et al,^40^ established a causal relationship between downregulation of BC200 RNA and oligodendroglial homeostasis, as targeted knockout of this gene leads to transcriptome and proteome changes in oligodendroglia-derived cell lines (MO3.13), ultimately resulting in their delayed differentiation and maturation.

Additionally, we performed an unbiased RNAseq analysis of small RNAs in lymphoblast cell lines from families F and H, carrying neighbouring variants in the bridge helix region. Comparison of DEGs in three affected individuals (F-I.1, F-II.1, H-II.1) in contrast to three unaffected family members (F-I.2, H-I.1, H-I.2) uniformly revealed a global decrease in the transcriptome of Pol III, compared to other polymerases. The downregulation was particularly pronounced for the tRNA pool, further demonstrating the Pol III dysfunction in patient cells. Notably, studies analysing transcriptome profiles of patients with biallelic Pol III-related disorders,^8,40,50,52^ *POLR3A*-HLD HeLa models,^40^ and a mouse *POLR3A*-HLD^44^ using different sequencing approaches also report global reduction in (primarily premature) tRNA levels. Unfortunately, there is limited consistency between the individual tRNA genes downregulated across the studies, including ours. Novel sequencing strategies that can capture mature tRNAs and unified disease models are required to test whether the different clinical Pol III entities result from differential tRNA expression in the affected neuronal populations.

Interestingly, other proteins involved in tRNA biogenesis have also been implicated in both hypomyelinating leukodystrophy and peripheral neuropathy, the prominent example being the aminoacyl-tRNA synthetase family. Biallelic pathogenic variants in *EPRS1*^53^ and *RARS1*,^54^ which encode aminoacyl-tRNA synthetases, as well as *AIMP1*^55^ and *AIMP2*,^56^ which encode structural components of the multi-tRNA synthetase complex, are known to cause hypomyelinating leukodystrophy. In contrast, monoallelic pathogenic variants in eight aminoacyl-tRNA synthetases cause peripheral neuropathy.^57–64^ Furthermore, in *AARS1,* monoallelic variants cause CMT disease,^59^ while biallelic variants have been associated with an infantile-onset encephalopathy featuring myoclonic seizures, reduced central myelination, and peripheral neuropathy^65^ – a phenotype encompassing features seen across both monoallelic and biallelic *POLR3A* and *POLR3B* disorders. Notably, ISR activation due to decreased tRNA supply restricted to the peripheral motor and sensory neurons of mouse models of *GARS1* and *YARS1* neuropathy has been reported.^66^ We did not observe increased ISR activation in EBV-transformed lymphoblasts of three *POLR3A*-neuropathy patients (F-I.1, F-II.1, H-II.1), however a neuron-specific effect cannot be excluded. Overall, our findings of a downregulated tRNA pool lend support to the emerging concept of disrupted tRNA metabolism as a driver of neurodegeneration in both the PNS and CNS.

The inheritance pattern of the neuropathy-associated variants is compatible with several disease mechanisms. Population data from gnomAD v4 show that *POLR3A* exhibits low constraint against protein-truncating variants (pLi=0; LOEUF=0.836), and no neurological phenotypes have been reported in heterozygous carriers of truncating variants. This, combined with our findings that POLR3A abundance and subcellular localization are not altered in patient-derived cells (Figure 3A, Supplementary Figure 4), suggest that haploinsufficiency is an unlikely mechanism underlying *POLR3A-*neuropathy. Additionally, the results from our interactome study suggest that defective subunit interactions leading to incomplete Pol III complex assembly is not the overarching disease mechanism. The identified monoallelic variants may act through an alternative dominant-negative mechanism, such as disrupted interactions with DNA template, or they may exert a neurotoxic gain-of-function effect the precise nature of which remains to be determined.

In conclusion, we demonstrate that monoallelic missense variants in *POLR3A* cause early-onset sensorimotor polyneuropathy, with or without additional tissue involvement. These findings expand the clinical spectrum of *POLR3A*-related disorders beyond the well-characterised biallelic phenotypes and warrant careful consideration or reinterpretation of heterozygous variants in this gene as part of the diagnostic evaluation of CMT patients. Our study provides critical insights into the allelic diversity of *POLR3A*-related disorders and establishes a framework for understanding the molecular pathomechanisms underlying these conditions. Further mechanistic studies are needed to understand why these monoallelic variants primarily affect the peripheral nervous system, which may reveal fundamental links between RNA metabolism and human disease.

## Acknowledgments

We are sincerely thankful to the participating families for their generous contribution to this study. We thank João Paulo Kitajima for assisting with variant querying at Mendelics Genomic Analysis and confirming maternity and paternity in Family D. We greatly appreciate the expert work carried out at the VIB Nucleomics Core, with special thanks to Stefaan Derveaux, Rekin’s Janky and Jolien Vandewinkel. We are grateful for the continued support by the BIOMINA Core Facility for Bioinformatics at the University of Antwerp.

