## Supplementary materials for "Monoallelic *POLR3A* variants cause a Pol III-related disorder characterized by peripheral neuropathy"

### A. Supplementary Methods and Results

#### I. Genetic analysis protocols for individual families

##### Family A

Genomic DNA was extracted from peripheral blood using the PureGene Kit (Qiagen) following the manufacturer's instructions. Genome sequencing (GS) for an affected individual (A-II.2) was outsourced to the Garvan Sequencing Platform. Paired-end sequencing reads of 150 bp were generated using the Illumina NovaSeq 6000 sequencing machine, with 30-fold average read depth. GS data processing was performed at the Centre for Population Genomics (CPG) following the DRAGEN GATK best practices pipeline. Reads were aligned to the hg38 reference genome using Dragmap (v1.3.0). Cohort-wide joint calling of single nucleotide variants (SNVs) and small insertion/deletion (indel) variants was performed using GATK HaplotypeCaller (v4.2.6.1) with '-dragen-mode' enabled. Sample sex and relatedness quality checks were performed using Somalier (v0.2.15).[1] Variants were annotated using VEP 105 and loaded into the web-based variant filtration platform, seqr.[2] A WGS search was conducted using the seqr platform for low minor allele frequency (<0.01) variants in CMT-related genes.[3] Segregation of candidate variants was conducted using Sanger sequencing (Supplementary Figure 1).

##### Family B and E

Exome sequencing (ES) for Family B was conducted at Beijing Genomics Institute (BGI). Exome capture utilized Agilent SureSelect v5 and samples were sequenced with 2x100 bp paired end reads on a HiSeq 4000 instrument. Reads were aligned to the GRCh37 reference genome using BWA[4] v0.7.12 and duplicates were marked with Picard[5] v1.90. An average read depth of 110X was achieved. Variants were called using GATK haplotype caller v3.4-46 and annotated through a custom annotation pipeline. Copy number variants (CNVs) were identified using two different algorithms: Conifer v0.2.2 for large CNVs and ExomeDepth for smaller CNVs. Parental data allowed for de. novo mutation (DNM) calling as previously described.[6] Known pathogenic short tandem repeats (STRs) were analysed with Expansion Hunter v3.1.2., using default settings. GS for Family E was performed by BGI on a BGISeq500 using a 2x100 bp paired-end reads, achieving a minimum median coverage per genome of 30X. Read mapping to the GRCh38 reference genome was conducted using Bwa-mem2 v2.2.1, with quality control assessed by Qualimap v2.2.1. Variant were called using GATK HaplotypeCaller v3.8. Structural variants (SVs) were identified using Manta Structural Variant Caller v1.1.0 (Illumina) and CNVs were detected using Canvas Copy Number Variant Caller v1.40.0 (Illumina). SNVs, SVs and CNVs were annotated via in-house developed pipeline. STRs were analysed using Expansion Hunter v3.1.2. with default settings. DNMs were called as previously described by Lelieveld et al.[6] and by using DeNovoCNN v1.1.0. Variants in the exome and genome data were interpreted in the same fashion. Candidate rare single nucleotide variants were identified by filtering for a gnomAD v.2.1 or v.3.1 frequency and an in-house frequency of <1%. CNVs and SVs were also filtered for a frequency of <1% using the 1,000 Genomes database and an internal database, requiring a minimum overlap of 90%. Additional annotations such as CADD score, SpliceAI, phyloP and gnomAD constraints, along with the patient's phenotype, were taken into account to identify potential disease-causing variants. For Family E, we screened for potential disease-causing variants associated with the seizures observed in this individual by using a diagnostic epilepsy panel. Orthogonal validation of the candidate POLR9A variants in the two families was conducted using Sanger sequencing (Supplementary Figure 1).

##### Family C and D

Target enrichment was performed on genomic DNA from the probands in families C and D using the Illumina DNA Prep with Enrichment and custom probes, followed by sequencing on the NovaSeq 6000 platform (Illumina). 99.57% and 99.14% of the exome kit was covered at least 10x and the average sampling depth was 105x and 68.47x in families C and D, respectively. Reads were aligned to the reference human genome hg38 using BWA[4] v.0.7.13-r1126. SNVs and small insertions and deletions were detected with the HaplotypeCaller algorithm in the GATK module.[7] The detected variants were annotated and filtered by the in-house annotation software at Mendelics. A filtering strategy was performed using gnomAD v.2.1 or v.3.1 frequency ≤0.5% for biallelic variants. The same threshold was used for the 1K Genomes database. For heterozygous variants, the threshold was 0% for both databases. Genotyping variants with score less than 100 were excluded. Synonymous and intronic variants that were not reported in ClinVar or nearby exon-intron junctions were excluded. Additional visual inspection was performed to reduce false positives. Orthogonal validation of the POLR9A variant c.2278C>T, p.(Arg760Cys) was conducted using Sanger sequencing for Family C and trio exome sequencing for Family D (Supplementary Figure 1). Paternity and maternity were confirmed in Family D using Somalier.[1] Additional common SNVs in.

POLR9A were manually curated in both probands to verify if the pathogenic variant occurred in different haplotypes (Supplementary Figure 2).

Family F

GS was performed on the proband and affected child at the University of Miami using an Illumina NovaSeq 6000 platform. The proband was sequenced to 40x coverage, while the affected child was sequenced to approximately 30x coverage. Reads were mapped to the GRCh37 reference genome using BWA[4] version 0.7.17, sorted by Sambamba, and had their optical duplicate reads marked by GATK's MarkDuplicates utility. From the bam file, SNVs and InDels were called using the HaplotypeCaller algorithm from GATK version 4.0.0.0. SVs were detected using Manta version 1.6.0[8] as implemented in Parliament2.[9] Outlier tandem repeats were detected using ExpansionHunter Denovo (EHDn)[10] and then genotyped alongside known pathogenic tandem repeats using ExpansionHunter version 5.0.0.[11] Processed variants were annotated and analysed using the GENESIS platform.[12] Segregation and orthogonal confirmation of the POLR9A variant was conducted using Sanger sequencing (Supplementary Figure 1).

Family G

Genomic DNA for the proband, unaffected parent and two unaffected siblings was extracted from blood using the Janus® chemagic MSM I automated system (Revvity). Proband DNA underwent testing on the nerve disease-targeted gene panel at Diagnostic Genomics, Pathwest (Perth, Australia), as previously described.[13] Illumina PCR-free GS of the proband, unaffected parents and two unaffected siblings was performed at the Kinghorn Centre for Clinical Genomics (KCCG; Darlinghurst, Australia). Paired-end sequencing reads of 150 bp were generated using the Illumina NovaSeq 6000 sequencing machine, with 30-fold average read depth. GS analysis was performed by the Centre for Population Genomics following the DRAGEN GATK best practices pipeline. Reads were aligned to the hg38 reference genome using Dragmap (v1.3.0). Cohort-wide joint calling of SNVs and small indels was performed using GATK HaplotypeCaller (v4.2.6.1) with "--dragen-mode" enabled. Sample gender and relatedness quality checks were performed using Somalier (v0.2.15).[1] Variants were annotated using VEP 105 and loaded into the web-based variant filtration platform, seqr.[2] Variant analysis was performed using the seqr platform, filtering for candidate variants according to 7) gnomAD v2 or v3 allele frequencies (<0.001), 8) mode of inheritance (autosomal recessive, X-linked, de novo or autosomal dominant), 9) variant type (for example, missense, nonsense, frameshift, canonical splicing, indel), and 0) in silico prediction scores (CADD, SpliceAI, PolyPhen2, SIFT, REVEL, FATHMM, VEST, MutationTaster, EIGEN). Candidate variants were further filtered according to manual investigations of gene function, gene and variant evolutionary constraint, and the patient's phenotype. Segregation and orthogonal confirmation of the POLR9A variant was conducted using manual curation of the genome sequencing reads on Integrative Genomic Viewer[14] (IGV) (Supplementary Figure 1).

As one parental DNA was not available for segregation studies, targeted long-read sequencing was performed using DNA samples from the proband and one parent to determine the phasing of the POLR9A variant. High molecular weight (HMW) DNA samples were transferred to the Garvan Sequencing Platform for targeted long-read sequencing analysis on Oxford Nanopore Technology (ONT) instruments. Prior to ONT library preparations, DNA was sheared to 20–25 kb fragment size using a MegaRuptor 3 instrument and visualized post-shearing on an Agilent FemtoPulse. Sequencing libraries were prepared from 3–5 μg of HMW DNA, using native library prep kit SQK-LSK114, according to manufacturer's instructions. Each library was loaded onto a R10.4.1 flow cell and sequenced on a PromethION device with live target selection/rejection executed by the ReadFish software package.[15] Detailed descriptions of software and hardware configurations used for Read-Fish are provided in a previous publication.[16] Samples were run for a maximum duration of 72 h, with nuclease flushes and library reloading performed at approximately 24- and 48-h timepoints for targeted sequencing runs, to maximize sequencing yield. Raw ONT sequencing data was converted to BLOW5 format[17] using slow5tools (v.0.3.0)[18] then base-called using Guppy (v6).[19] Resulting FASTQ files were aligned to the hg38 reference genome using minimap2 (v2.14-r883).[20] Variants were called using clair3,[21] phased using WhatsHap[22] and visualised using the Integrative Genomics Viewer (IGV, v2.17.3).[14] Visualisation of reads at the POLR9A variant (c.2558C>T, p.Ala853Val) locus showed the variant to be on the parental allele that could not be sequenced (data not shown). Thus, de novo inheritance of the POLR9A variant was not able to be confirmed for the proband.

Family H

Genomic DNA was extracted from peripheral blood mononuclear cells according to standard protocols. ES was performed in proband H-II.1 using SeqCap EZ Exome Probes v3.0 kit (Roche Holding AG, Basel, Switzerland) for capture. Then, 150 bp exome paired-end sequencing was run on NextSeq 150 platform (Illumina, San Diego, CA). Read alignment to hg38 reference human genome was done with Burrows-Wheeler algorithm (version 0.7.15-r1140) and genomic analysis toolkit (GATK) (version 4.1.8.1) for variant calling. Long-read genome sequencing (lrGS) of the patient H-II.1 and the parents H-I.1 and H-I.2 was performed using the sequencing platform from Oxford Nanopore Technologies. Libraries of sizes ranging from 20-25 kb were generated and sequenced on a PromethION device using R10.4.1 flow cells. Reads were called with Guppy[19] and aligned to human genome reference hg38 with minimap2.[20] SVs were called with Sniffles2[23] and cuteSV,[24] and SNVs with Clair3[21] and Longshot.[25] Variant files from exome and lrGS were annotated with GenomeComb.[26] Variant prioritization was based on allele frequency using gnomAD v2 and v3 (<1%), and an internal cohort of 265 lrGS, predicted impact of the variant on the gene product according to in.silico.prediction tools (AlphaMissense, CADD, SpliceAI), patient phenotype and gene constraint metrics. All shortlisted variants were reviewed using IGV.[14] Known pathogenic expansions were excluded (STRchive)[27] by visual inspection of lrGS on IGV. Segregation and orthogonal confirmation of the POLR9A variant was conducted using Sanger sequencing (Supplementary Figure 1).

#### II. In.silico.analyses

We screened the Human Gene Mutation Database (HGMD, https://www.hgmd.cf.ac.uk/), Leiden Open Variation Database (LOVD, https://www.lovd.nl/), ClinVar database (https://www.ncbi.nlm.nih.gov/clinvar/) and the available literature to identify pathogenic missense variants in POLR9A. associated with a disease phenotype. Additionally, we screened the gnomAD database (https://gnomad.broadinstitute.org/) to look for missense variants that were observed in homozygosity in at least one individual in gnomAD v3 or observed in more than 10 controls in gnomAD v2 which we considered "potentially benign". We then compiled this list of missense variants in POLR9A.and curated it with information including the phenotype associated with each variant, its zygosity, number of heterozygous individuals reported in gnomAD v3, original and variant amino acid category, AlphaMissense pathogenicity score,[28] MetaDome mutational tolerance score,[29] evolutionary conservation percentage of wild-type and mutant residues provided by Protein Data Bank in Europe Knowledge Base (PDBe-KB),[30] relative surface accessibility of wild-type residues as calculated by FreeSASA software[31] v.2.0.324, position of the residue within POLR3A protein domains, ClinVar accession number and significance (if reported), and the PMID accession number of the original manuscript reporting each variant when present. This list contains 89 missense variants in POLR9A with 71 variants associated with biallelic neurological phenotypes, three variants associated with susceptibility to severe reactions to Varicella zoster virus infections, seven variants associated with peripheral neuropathy with variable central nervous system involvement reported in this study, and eight "potentially benign" missense variants. Strikingly, in such a large protein, we were able to identify very few "potentially benign" variants which is also corroborated by the high constraint score for this gene in gnomAD v4.1.0 (missense z-score=3.68). We analysed AlphaMissense pathogenicity scores of different groups of neurological phenotypes and observed that missense variants associated with demyelinating peripheral neuropathy had an average score of 0.97, while missense variants associated with recessive disorders (HLD, WRS, or spastic ataxia/spastic paraparesis) had an average pathogenicity score of 0.87. This difference between the average pathogenicity scores was not statistically significant; however, the pathogenicity scores of potentially benign missense variants were significantly lower (Supplementary Figure 3B).

We used PDB entry 7AE3 to calculate relative surface accessibility (RSA) scores of individual residues using the FreeSASA software[31] v.2.0.324 with default parameters. When modelling the entire Pol III complex, all residues had low RSA scores indicating that the affected residues are buried inside the enzymatic complex and not exposed to the surface of the polymerase. Moreover, when the nucleic acids are modelled into the Pol III complex, the RSA score of p.Arg360 is further reduced, indicating that this residue interacts with nucleic acids within the complex (Supplementary Figure 3A). This was also evident in the cryo-EM structure of human Pol III which indicates p.Arg360 contacting the template DNA strand at the transcription bubble.[32]

Using the variants clustering to the funnel domain and the bridge helix region of POLR3A, we also assessed the alterations of size and physicochemical characteristics of amino acid residues impacted by variants in monoallelic vs. biallelic neurological disorders. Size categories were defined as tiny (with maximum two heavy atoms in the side chain: Ala, Cys, Gly, Ser), medium small (three heavy atoms in the side chain: Asn, Asp, Pro, Thr, Val), and large (minimum four heavy atoms in the side chain: Arg, Gln, Glu, His, Ile, Leu, Lys, Met, Phe, Trp, Tyr). Chemical categories of amino acids are defined as positively charged, negatively charged, polar, hydrophobic, and special (cysteine, glycine, proline are defined as special cases). Comparison of residue size and chemistry between original and variant amino acids show no special clustering to certain groups in different diseases, however, neuropathy-associated variants in general have more drastic alterations than recessive disease-causing variants (Supplementary Figure 3C).

#### III. Proteomics analyses

Cell culture

Plasmid vector construct generation and sequence verification was done by Genscript Biotech (APAC Division, Singapore). Variants were introduced using site directed mutagenesis into the cDNA of POLR3A and subsequently cloned into a pCMV-3Tag-3a expression vector with a C-terminal FLAG epitope (C-3*DYKDDDDK). Tested variants are: c.1024G>A, p.Val342Ile; c.1078A>G, p.Arg360Gly; c.1724A>T, p.Lys575Met; c.2270C>T, p.Ser757Phe; c.2357A>G, p.Lys786Arg; c.2548C>A, p.His850Asn; c.2558C>T, p.Ala853Val; c.2560G>A, p.Gly854Ser. Among these, p.Val342Ile and p.Lys575Met are used in data analysis as benign substitutions since p.Lys575Met has high allele count in gnomAD v4 and is classified as benign/likely benign on ClinVar by multiple submitters (VCV000301068.21) and the p.Val342Ile variant has a high heterozygote count in gnomAD v4 and is in close proximity to the p.Arg360Gly candidate variant.

Human embryonic kidney (HEK293) cells were grown and maintained in DMEM medium with high-glucose (4500 mg/L) (supplemented with 10% foetal bovine serum (Performance plus, USA, Gibco), 2mM L-glutamine, and 100 U/mL penicillin and 100µg/mL streptomycin). Cells were then plated at a density of $1.6x10^7$ cells per 15-cm plates and were grown overnight. Plasmid vector constructs containing either the wild-type or mutant POLR9A were transfected into cells using Lipofectamine3000 (Thermofisher) according to manufacturer's instructions. Untransfected cells were grown as usual and were subjected to the addition of OPTI-MEM instead of plasmids at the transfection step. Transfected cells were incubated at 37°C for 24 h. Cells were harvested using a cell scraper in growth media in a 50mL Falcon tube. Cells were pelleted at room temperature (300 x g for 5 mins), before decanting the supernatant and resuspending in 1 mL ice-cold PBS. Resuspended cells were transferred to a 1.5mL tube and pelleted at 4°C using a centrifuge (500 x g for 5 mins). This step was repeated twice. Mass of the cell pellet was determined (approximately 250-500mg of dry cell pellet). Cells were snap frozen in dry ice after harvesting. The total cell amount from two 15 cm plates was considered one biological replicate.

Affinity purification

Transfected and untransfected cell pellets were solubilised at a 1:4 pellet weight to volume ratio in a digitonin-containing solubilisation buffer (20 mM Tris-Cl pH 7.4, 50 mM NaCl, 10% v/v glycerol, 0.5 mM EDTA, 1% w/v digitonin, and 25 U benzonase), and incubated at 4°C for 30 min. Lysed samples were clarified by centrifugation at 20,000 x g for 5 min at 4°C, and protein concentration was determined using the Pierce BCA Protein Assay Kit (ThermoFisher). 1 mg of total protein was aliquoted for each replicate, and the total volume was brought up to 500 µl with solubilisation buffer. Samples underwent affinity enrichment to capture FLAG-interacting proteins, as previously described[33] with modifications detailed below. Affinity purification was performed in technical triplicate for each sample.

Pierce Spin Columns (ThermoFisher) were loaded with 40 µl anti-FLAG MS2 affinity resin and equilibrated with wash buffer (20 mM Tris-Cl pH 7.4, 60 mM NaCl, 10% v/v glycerol, 0.5 mM EDTA, 0.1% w/v digitonin). Samples were loaded onto spin columns containing anti-FLAG resin beads and incubated at 4°C for 2 h while rotating. Samples were washed 15 times with 500 µl wash buffer, followed by incubation with 50 µl wash buffer containing 100 µg/ml FLAG peptide (Sigma) at 4°C for 1 hour with rotation to elute FLAG bound proteins. Eluted proteins were collected by centrifugation into a microcentrifuge tube, and an additional 50 µl of wash buffer without FLAG peptide was added and centrifuged into the same tube. Eluted proteins were prepared for mass spectrometry by adding a final composition of 5% SDS and 50 mM triethylammonium bicarbonate (TEAB) pH 8.5, followed by processing using S-trap spin Columns (Protifi) according to the manufacturer's instructions. Proteins were digested with 0.5 µg trypsin at 37°C overnight and eluted peptides were dried down using a CentriVap Benchtop Vacuum Concentrator (Labconoco).

Mass spectrometry

Samples were reconstituted in 0.1% trifluoracetic acid (TFA) and 2% acetonitrile (ACN) for analysis by liquid chromatography (LC)-tandem mass spectrometry (MS/MS) on a Q Exactive HF-X mass spectrometer (Thermo Fisher Scientific) coupled with an Ultimate 3000 RSLC nanoHPLC (Dionex Ultimate 3000), operating on data-dependent acquisition mode. The samples were loaded on an Acclaim Pepmap nano-trap column (Dionex-C18, 2 cm) at an isocratic flow of 5 µl/min of 2% ACN containing 0.1% formic acid (FA) for 5 min applied before switching to a µPAC™ Neo HPLC analytical column (Neo-C18, 50 cm). The separation of peptides was performed over a 65-minute gradient of solvent A (5% dimethyl sulphoxide (DMSO), 0.1% FA) and solvent B (5% DMSO, 100% ACN, 0.1% FA). The flow gradient was (i) 0–6 min at 3% solvent B, (ii) 6-35 min, 3-23% solvent B, (iii) 35-45 min, 23-40% solvent B, (iv) 45-50 min, 40-80% solvent B, (v) 50-55 min, 80% solvent B, (vi) 55-56 min, 80-5% solvent B, and equilibrated at 5% solvent B for 10 min before the next sample injection. Briefly, full MS spectra were acquired in positive mode at 120,000 resolution, an automatic gain control (AGC) target of $3e^6$ and a maximum injection time (IT) of 50 ms. The 12 most intense peptide ions with a z≥2 and an intensity threshold of $1.8e^5$ were subjected to MS/MS using high-intensity collision dissociation. For MS2, the isolation window was 1.2 m/z and precursors were fragmented using a normalised collision energy of 30%. The resolution was 15,000 with AGC target of $1e^5$ and maximum IT of 22 ms. Dynamic exclusion was set at 20 s.

Raw files were analysed using the MaxQuant platform (version 1.6.10.43)[34] and searched against the UniProt human protein database (exported March 2021). Default LFQ parameters were applied, with the LFQ min. ratio count and Label min. ratio count set to 2. Subsequent analysis was performed using Perseus (version 1.6.15.0).[35] LFQ intensities were imported from the proteinGroups.txt output and $log_2$ transformed. Proteins labelled as "Only identified by site", "Reverse", or "Potential contaminant" were removed. Each comparison of the experimental samples (Flag-POLR3B Mutant X) cell line was compared to the untransfected control independently. Proteins quantified in at least 2 of the experimental sample replicates were included, and missing values in the untransfected control were imputed for based on the normal distribution. A two-sided t test was performed and visualised using the scatter-plot function, with significance set to p_value – 0.05 ($-log_{10}$ = 1.301) and fold-change +2 ($log_2$ = 1).

It is important to note that the comparisons between the mutant and wild-type POLR3A are only based on subunits that were detected in both the wild-type and mutant conditions, as approximately half of the 17 Pol III subunits were not detected by the assay.

#### IV. Transcriptomics analyses

RNA extraction from the two affected individuals (F.I-1 and F.II-1), one unaffected individual (F.I-2) from Family F and one affected individual (H.II-1) and two unaffected individuals (H.I-1 and H.I-2) from Family H was performed using the Qiagen miRNeasy kit with a modified extraction protocol designed to isolate small RNA (<200 nucleotides) enriched fragment. RNA concentration and purity were measured using a NanoDrop spectrophotometer and RNA integrity and concentration were assessed using a BioAnalyzer Nano kit (Agilent, Santa Clara, CA, USA). 100 ng RNA per sample was used as input. Two small RNAs (70 and 94 nucleotides) were synthesized and spiked in into the libraries at the beginning of the protocol alongside the standard External RNA Controls Consortium (ERCC) spike-in mix.

Small RNA libraries were prepared according to the Illumina Stranded Total RNA preparation protocol with ligation using the Ribo-Zero Plus kit, however depletion steps were excluded, and the clean-ups were less stringent (1.2X) on the adapter-ligated fragments and on the dual-indexed libraries. The small RNA samples were fragmented and used in a first strand reverse transcription reaction using random primers and Actinomycin D and subsequently converted into double-stranded cDNA in a second strand cDNA synthesis reaction using dUTP to achieve strand specificity. The cDNA fragments were extended with a single 'A' base to the 3' ends of the blunt-ended cDNA fragments after which pre-index anchors were ligated preparing the fragments for dual indexing. Anchor-ligated fragments were then purified using magnetic beads. Finally, enrichment PCR was carried out to enrich anchor-ligated DNA fragments and to add indexes and primer sequences for cluster generation. Purified dual-indexed sequencing libraries of each sample were equimolarly pooled and a double-sided clean-up was performed on the pool to remove as much ERCC mix and primer dimers as possible. Purified, dual-indexed libraries were sequenced on Element Biosciences AVITI instrument following the parameters AVITI 150 cycles Cloudbreak Freestyle sequencing kit, single-end reads (151-10-10-0) with 2% PhiX at the VIB Nucleomics Core.

The sequencing reached an assigned yield of 79.690 Gb. After demultiplexing at the VIB Nucleomics core, we obtained 535.098M pass-filter reads, corresponding to 584.609M raw reads in six samples analysed. 87.7% of the reads passed the Q30 quality threshold with 74.3% of reads passing the Q40 threshold.

Quality control of Fastq files were performed using FastQC v0.12.0.[36] Low quality bases at the ends of raw reads and adapter dimer contamination were removed. Cutadapt[37] was used to clean and trim small RNA samples, keeping the reads with a minimum read length of 18 after trimming. Sequenced reads were mapped to human reference genome hg38 including tRNA genes extracted from the tRNA database[38] using STAR v.2.7.10b allowing all multimapping reads. RSeQC[39] was used to assess tRNA gene body coverage. Gene counts were extracted using featureCounts in three successive analyses using 7). all mapped reads, 8) only primary alignments, and 9) only uniquely mapped reads. Library normalization and differential expression analysis were performed with the DESeq2 pipeline.[40] We verified that the results overlapped with all three protocols and provided the data belonging to primary alignments in the original display items.

DESeq2 analysis comparing the affected individuals with unaffected family members on the primary alignments resulted in 187 differentially expressed genes (p-adjusted<0.05) among which 23 being individual tRNA genes.

#### B. Supplementary Figures

Supplementary Figure 1: Comparative sequence alignment of RNA Polymerase subunits and validation of POLR9A.variants in the families. A. Multiple sequence alignment of the largest subunits of human RNA polymerase I, II and III, encoded by POLR7A, POLR8A.and POLR9A, respectively, along with the corresponding POLR3A subunits from various species, indicating identical or highly conserved residues (dark blue) and moderately conserved residues (light blue). The highly conserved molecular hinges made up of glycine residues in the bridge helix region are shown in pink as $H_C$ (carboxyl-terminal hinge) and $H_N$ (amino-terminal hinge). Hs: Homo.sapiens; Mm: Mus.musculus; Dr: Danio.rerio; Dm: Drosophila.melanogaster; Ce: Caenorhabditis.elegans; Sc: Saccharomyces. cerevisiae. B. Sanger sequencing electropherograms in families A, B, C, E, F and H for the validation of the POLR9A.variants. C. Screenshot from IGV showing the sequencing data for the variant in POLR9A.for families D and G.

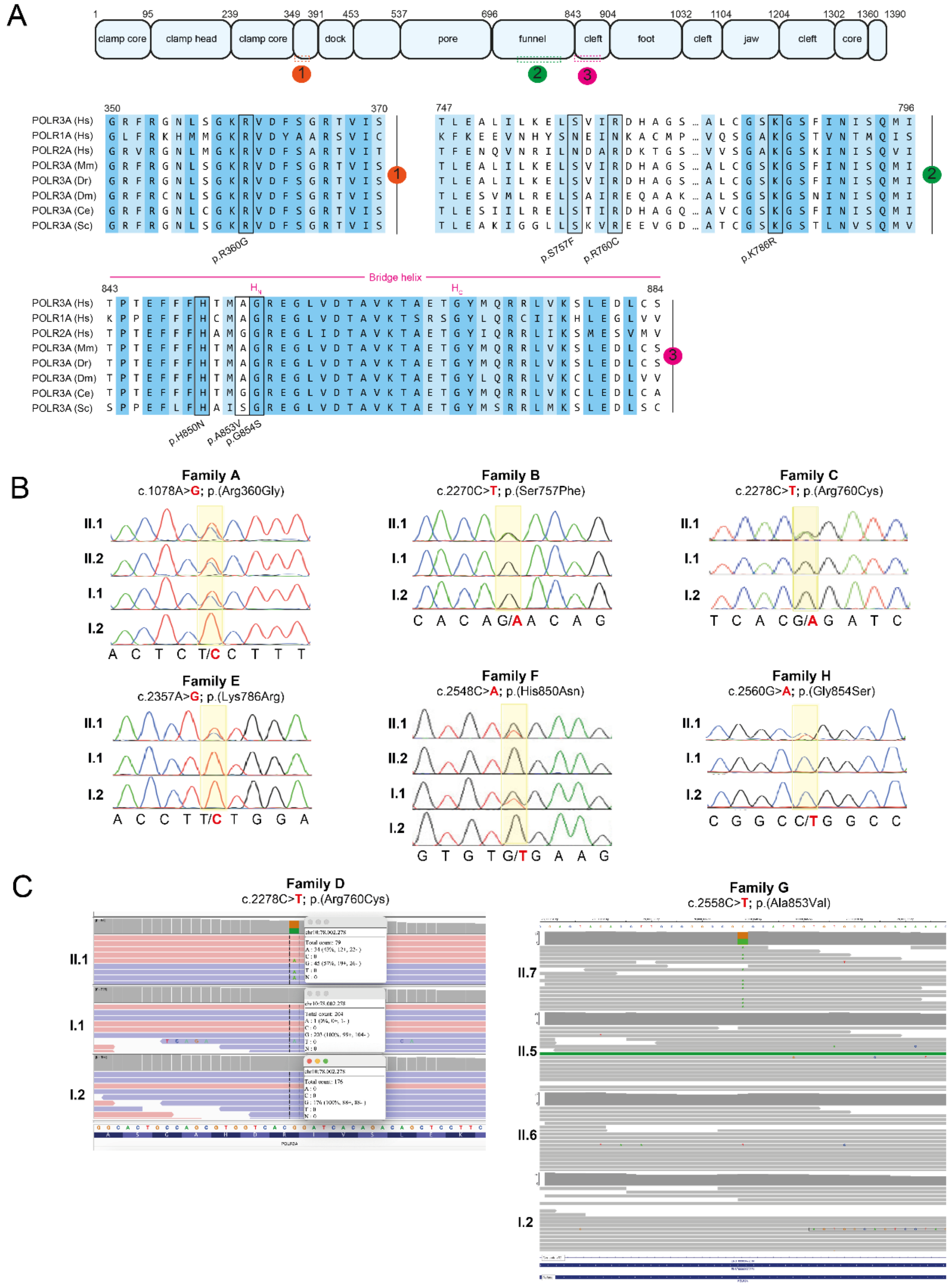

Supplementary Figure 2: Haplotype analysis in patients C-II.1 and D-II.1. A. Table with SNVs residing in POLR9A. and corresponding zygosity for each individual. Notably, variant chr10-78002270-G-A is in cis with the pathogenic variant chr10-78002270-G-A in individual C-II.1 and absent in individual D-II.1. In addition, two variants are homozygous in C-II.1 and absent in D-II.1. The pathogenic variant is highlighted in bold; B. Screenshots of cram files for both patients show multiple variants in POLR9A in individual C-II.1 that are absent in individual D-II.1, demonstrating the pathogenic de.novo variant arose on distinct haplotypes.

A

| Variant | C-II.1 | D-II.1 | HGVSp/ HGVSc |
|---|---|---|---|
| chr10-78009910-T-A | absent | heterozygous | p.Lys575Met |
| **chr10-78002278-G-A** | **heterozygous** | **heterozygous** | **p.Arg760Cys** |
| chr10-78002270-G-A | heterozygous | absent | p.His762His |
| chr10-78001060-A-G | heterozygous | absent | p.Cys798Cys |
| chr10-78000928-T-G | homozygous | absent | c.2478+48A>C |
| chr10-77986031-A-G | homozygous | absent | c.2988+42T>C |

B

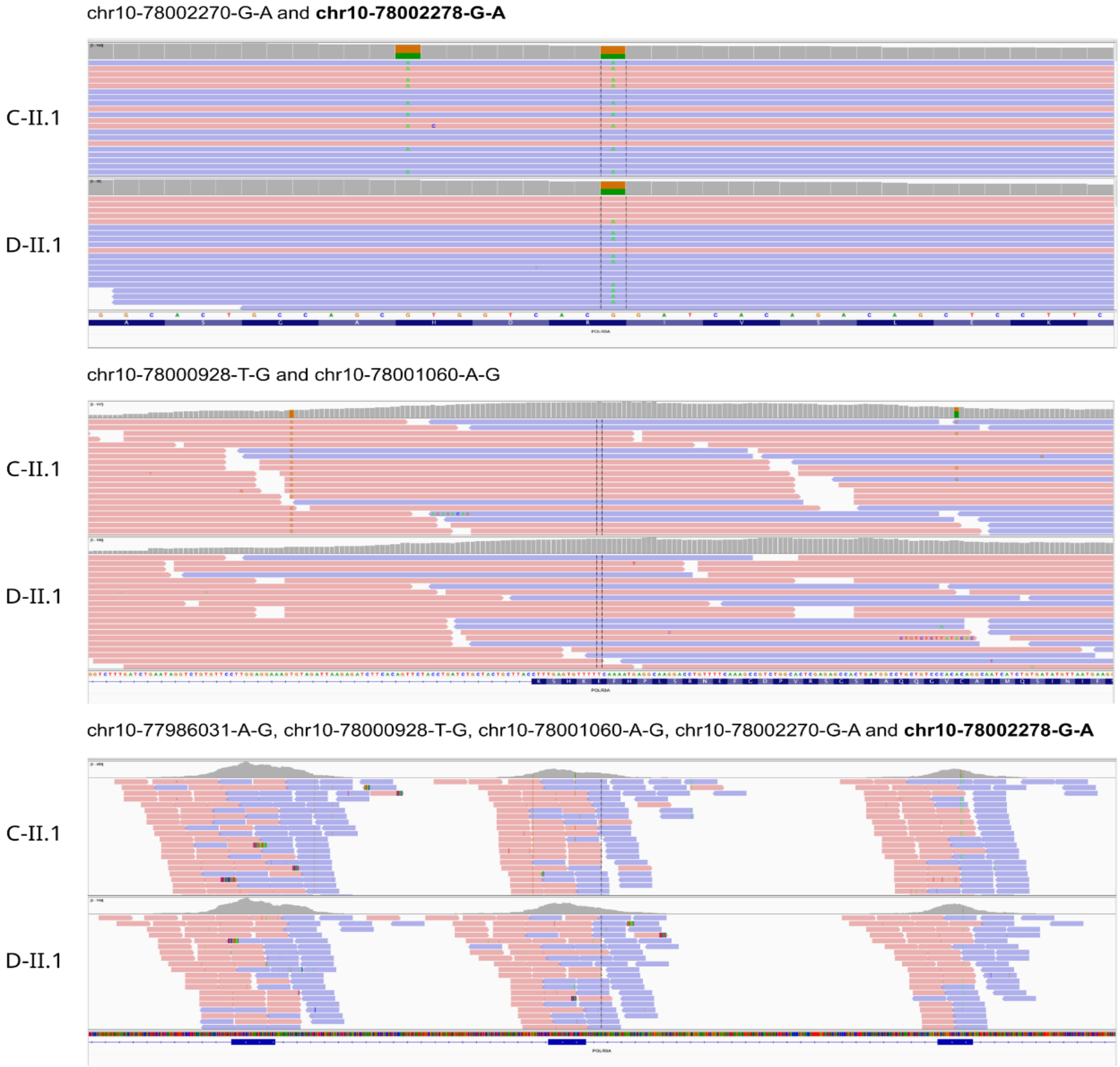

Supplementary Figure 3: Additional in. silico findings. A. Relative surface accessibility (RSA) percentages of the amino acid residues of POLR3A affected by the neuropathy-associated substitutions are shown for isolated POLR3A protein, the assembled polymerase complex and the complete polymerase model including nucleic acids (Nas). Lower RSA values indicate reduced solvent exposure reflecting the residues are more buried within the structural context of the modelled molecule(s). B. Violin plot comparing the distribution of AlphaMissense pathogenicity scores for POLR9A missense variants associated with monoallelic and biallelic disorders, and likely benign variants with relatively high population frequencies in gnomAD. Mean pathogenicity scores do not significantly differ between monoallelic and biallelic disorders; however, both groups have significantly higher pathogenicity scores relative to "benign" variants (****:p<0.0001, one-way ANOVA with multiple comparisons). C-D. Percentage distribution of alteration types based on the size and physicochemical characteristics of amino acid residues in the funnel domain and bridge helix region of POLR3A impacted by peripheral neuropathy-associated and biallelic disorder-causing substitutions, respectively. Amino acid size categories are defined as tiny (maximum two heavy atoms in the side chain: Ala, Cys, Gly, Ser), medium small (three heavy atoms in the side chain: Asn, Asp, Pro, Thr, Val), and large (minimum four heavy atoms in the side chain: Arg, Gln, Glu, His, Ile, Leu, Lys, Met, Phe, Trp, Tyr). Chemical classifications of amino acids are defined as positively charged, negatively charged, polar, hydrophobic, and special (cysteine, glycine, proline are defined as special cases). While no specific clustering of residue changes was observed across different diseases, neuropathy-associated variants generally involved more substantial changes in size and chemical properties compared to the recessive disease-causing variants.

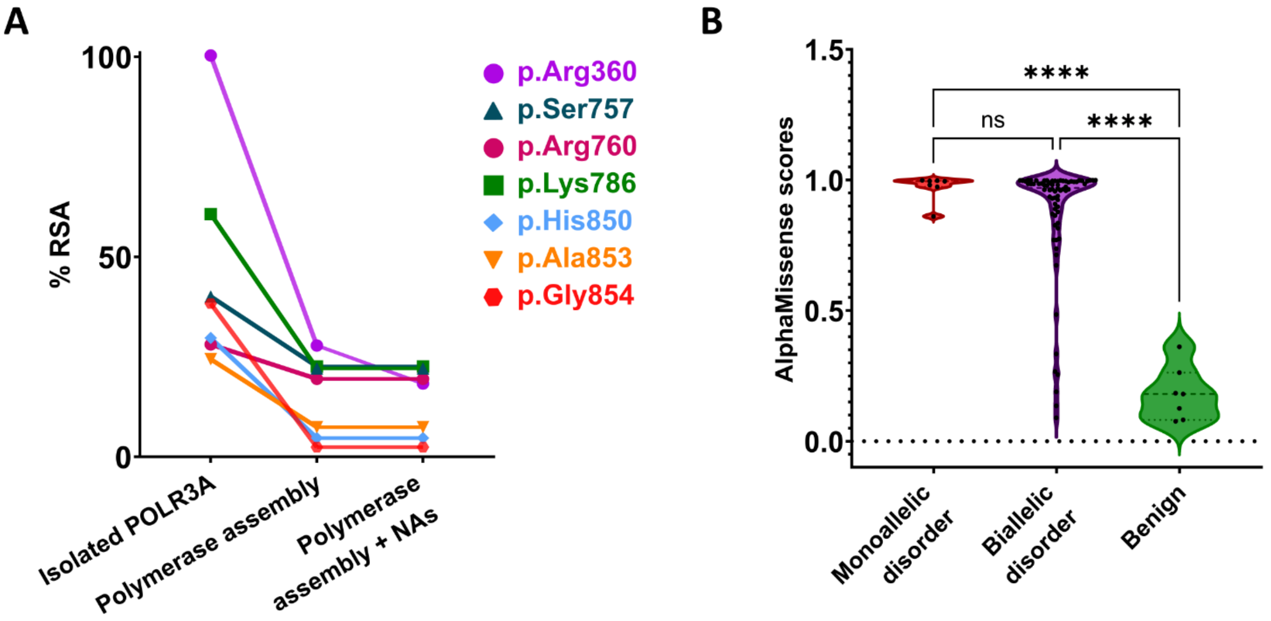

C

| Peripheral neuropathy-causing substitutions | Original chemistry | Altered chemistry |
|---|---|---|
| Original size | 17% | 17% |
| Altered size | 17% | 50% |

D

| Recessive disorder-causing substitutions | Original chemistry | Altered chemistry |
|---|---|---|
| Original size | 33% | 27% |
| Altered size | 7% | 33% |

Supplementary Figure 4: Immunofluorescence staining for POLR3A in skin-derived human fibroblasts comparing a control cell line with individual E-II.1 harbouring the p.(Lys786Arg) variant. In both cell lines, POLR3A showed a similar distribution pattern, with predominant nuclear localization.

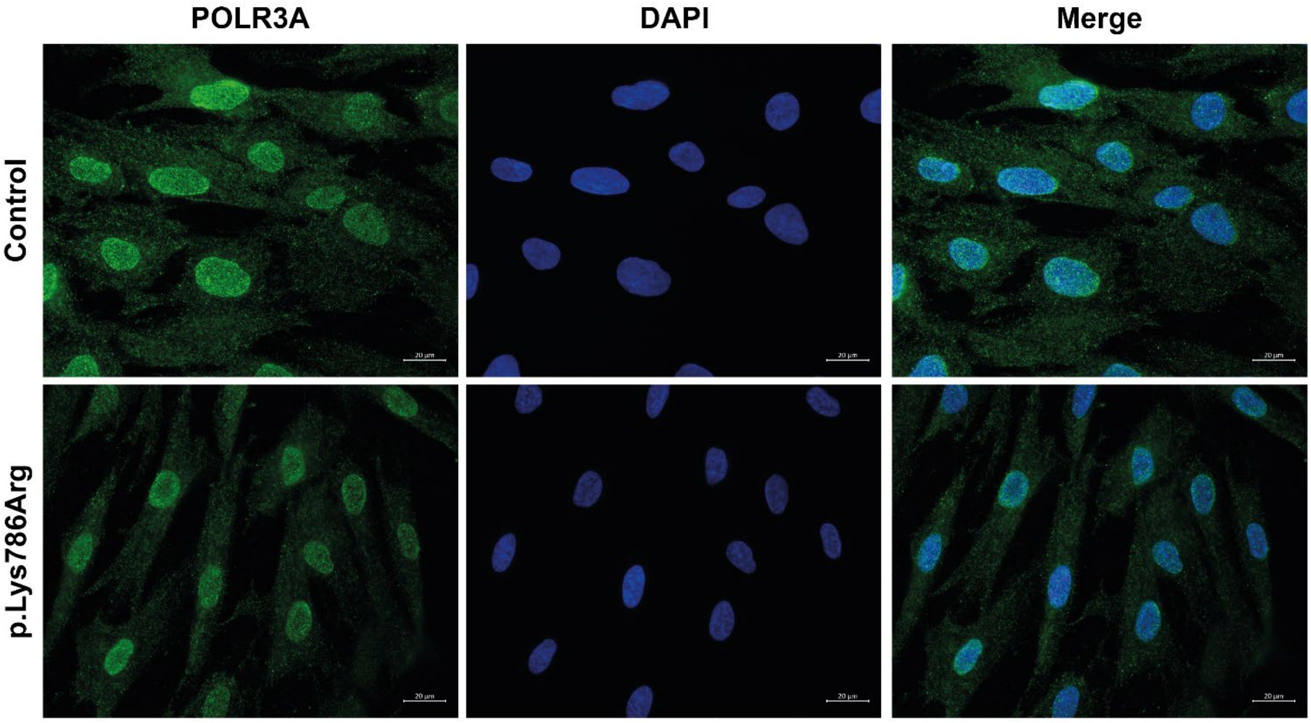

Supplementary Figure 5. Additional findings from the affinity purification coupled with mass spectrometry assay of mutant POLR3A. A: Volcano plots showing $\log_2$ fold-change between wild-type (WT) and untransfected (UT) or POLR3A mutant and UT, against the $-\log_{10}$ p-value obtained via a two-tailed t-test. B: Volcano plot showing t-test comparison of mutant POLR3A (patient or benign variants) directly with WT POLR3A. To compare mutant POLR3A with WT POLR3A, all values of all proteins in each replicate were normalised to POLR3A abundance within that replicate, followed by a t-test. Results were filtered to only show proteins which were significantly enriched ($p < 0.05$, $\log_2$fold-change > 1) from A. All Pol III subunits were included in B (dark blue) regardless of significance. For A and B, Overexpressed POLR3A is labelled in red, while components of the RNA Pol III enzyme complex are labelled in blue. Vertical line indicates a log2 fold-change > 1, while the horizontal line represents $p < 0.05$. N=3 replicates for all WT and mutant samples.

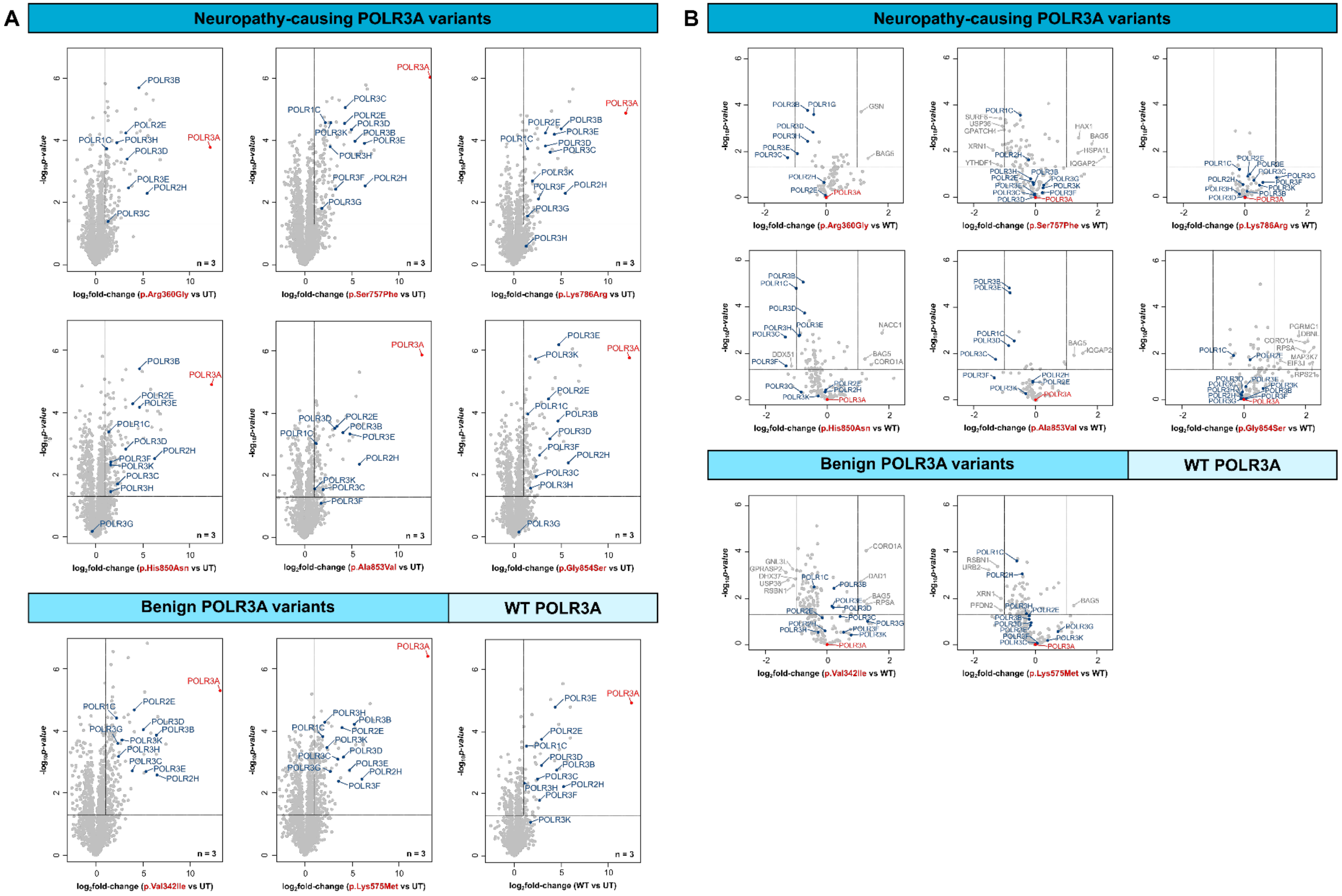

Supplementary Figure 6: qRT-PCR expression analysis of transcript tRNA-Asn-GTT-10-1 across the neuropathy cohort. Relative transcript levels were normalized to three housekeeping genes (PSMB❷, SDHA, and COPS❸A). No significant changes were observed in patients with a substitution in the DNA-binding residue (Family A). In contrast, patients with funnel domain substitutions (B-II.1 and E-II.1) showed upregulation of tRNA-Asn-GTT-10-1, while individuals with bridge helix substitutions (F-I.1, F-II.1, G-II.7, and H-II.1) showed significant downregulation of this transcript. Unpaired t-test with Welch's correction is used for statistical analysis. Significance levels are indicated as follows: ns: not significant, *:p<0.05, **:p<0.01, ***:p<0.001, ****:p<0.0001.

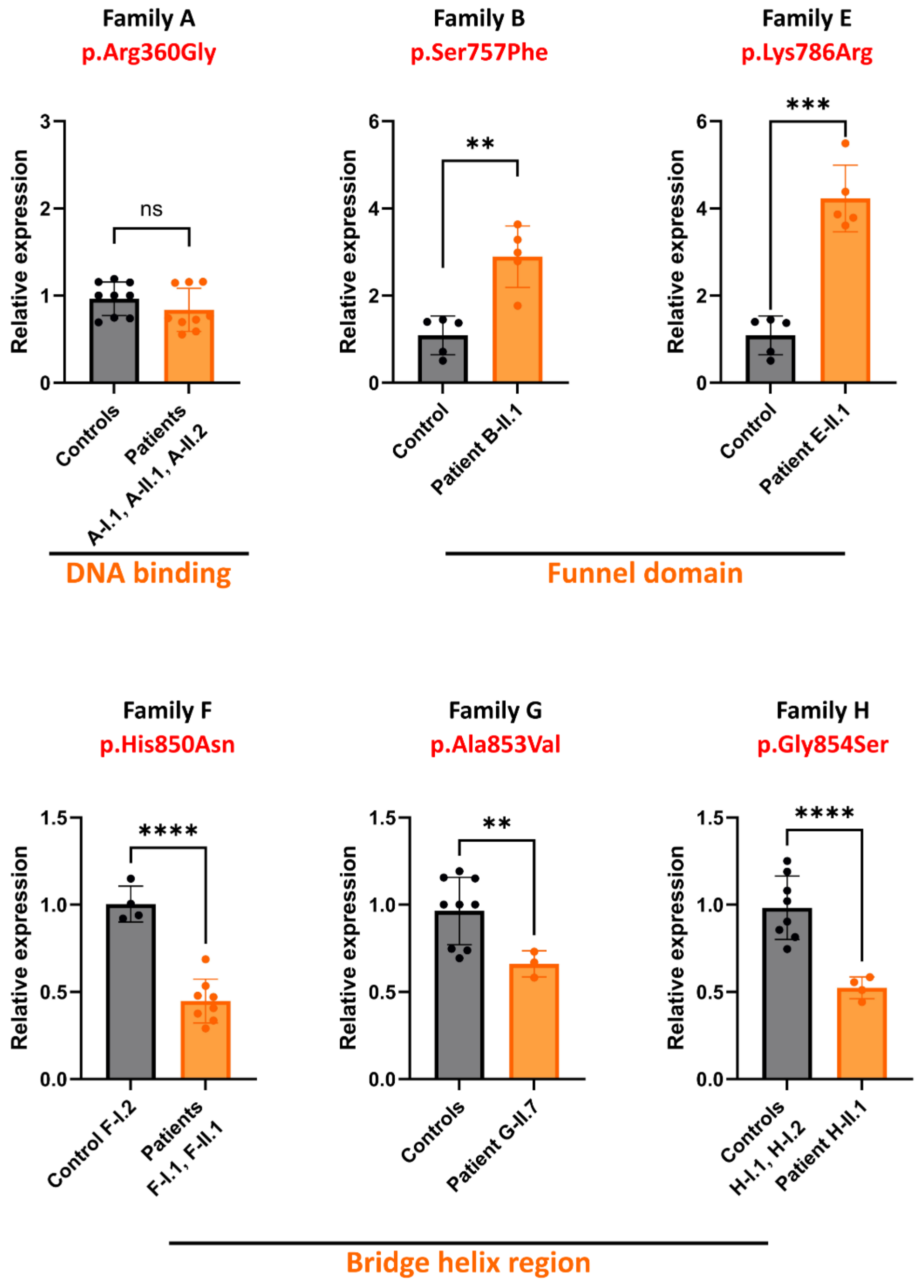

Supplementary Figure 7: Western blot analysis of integrated stress response (ISR) pathway markers in Family F and H. A. Representative western blot images showing expression levels of the major ISR components (top) and the full blot image after puromycin treatment of cells (bottom). B. Quantification of relative expression of the blots from panel A normalized to α-tubulin abundance. Bar graphs display relative expression in patient-derived EBV-transformed lymphoblasts versus unaffected family members for each marker. No significant differences were observed between groups (unpaired t-test; p-values indicated above each comparison).

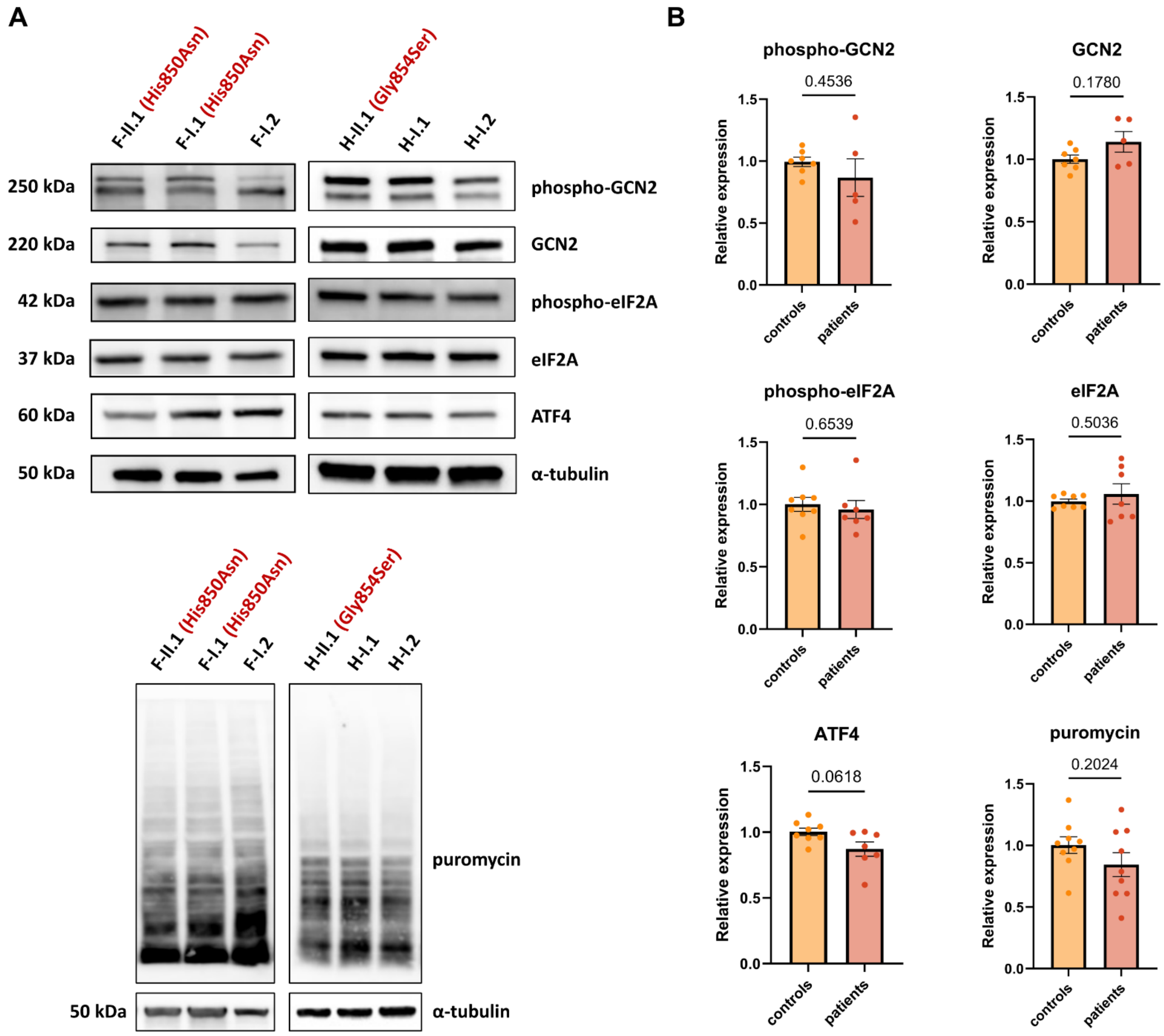

#### C. Supplementary Tables

Supplementary Table 1: Oligonucleotide sequences used in the study

| Primer name | Sequence (5'-3') | Purpose | Reference |
|---|---|---|---|
| POLR3A_ex8_F | AGTGTCTGTCCAGCTTCTC | Sanger validation | This study |
| POLR3A_ex8_R | TGGCACAGCTACCTCATCAA | | |
| POLR3A_ex10_F | AGTCCACTGTTTAGCACTGAAC | Sanger validation | This study |
| POLR3A_ex10_R | ACTTGGTGAGAGAACTTTGCT | | |
| POLR3A_ex19_F | ATCGTGGGCCTTGAACTTTT | Sanger validation | This study |
| POLR3A_ex19_R | GTCCTGCTCTGTTGCTAATGG | | |
| POLR3A_ex17_F | TGGAGCCAAACAGCCTGTAC | Sanger validation | This study |
| POLR3A_ex17_R | CACAGAGACAATGAGGACAACC | | |
| POLR3A_ex27_F | TGTGTTGCCATGAACTGAGC | Sanger validation | This study |
| POLR3A_ex27_R | TCTAACAGACTAACTCCTTGGGG | | |
| 7SL_RNA_F | GGAGTTCTGGGCTGTAGTGC | qRT-PCR | [41] |
| 7SL_RNA_R | TTTGACCTGCTCCGTTTCCG | | |
| 7SK_RNA_F | AGAGGACGACCATCCCCGAT | qRT-PCR | [42] |
| 7SK_RNA_R | TGGAAGCTTGACTACCCTACGT | | |
| 5S_rRNA_F | GCCATACCACCCTGAACGC | qRT-PCR | [41] |
| 5S_rRNA_R | TATTCCCAGGCGGTCTCCC | | |
| BC200_F | ATATAGCGAGACCCCGTTCT | qRT-PCR | This study |
| BC200_R | TGCTTTGAGGGAAGTTACGC | | |
| tRNA_Leu_CAA_1_2_F | CTCAAGCTTGGCTTCCTCGT | qRT-PCR | [43] |
| tRNA_Leu_CAA_1_2_R | GAACCCACGCCTCCATTG | | |
| tRNA_Ala_TGC_1_1_F | GAACCCGGGACCTCATACAT | qRT-PCR | [43] |
| tRNA_Ala_TGC_1_1_R | GGGGGTGTAGCTCAGTGGTA | | |
| tRNA_Tyr_GTA_8_1_F | AGCGGAGGACTGTAGGTTCA | qRT-PCR | [43] |
| tRNA_Tyr_GTA_8_1_R | GATTCGAACCAGCGACCTAA | | |
| tRNA_Gly_TCC_4_1_F | GTTGGTGGTATAGTGGTGAGCA | qRT-PCR | [42] |
| tRNA_Gly_TCC_4_1_R | TGCGTTGGGCGGGAATC | | |
| tRNA_iMet_CAT_1_F | GCAGCGGAAGCGTGCT | qRT-PCR | [42] |
| tRNA_iMet_CAT_1_R | AGCAGAGGATGGTTTCGATCC | | |
| POLR3A_F | CTTCCAAGGCCATCAGCACT | qRT-PCR | This study |
| POLR3A_R | ATTCTCCCTTTCACGAGGCG | | |
| POLR3B_F | CTTCGTGAGCAGCATGGAC | qRT-PCR | This study |
| POLR3B_R | CAGCCTCCATTTTTCCTCTACA | | |
| tRNA-Asn-GTT-10-1_F | CTCTGTGGCGCAATCGGCT | qRT-PCR | This study |
| tRNA-Asn-GTT-10-1_R | GTGGGTTCGAACCGCCAA | | |
| tRNA-Asn-GTT-4-1_F | TCTGTGGCGCAATCGGTT | qRT-PCR | This study |
| tRNA-Asn-GTT-4-1_R | GAACCACCAATCTTTCGGTT | | |
| tRNA-Asn-GTT-9-2_F | CCGGGTGGGCTCGAACCACTAA | qRT-PCR | This study |
| tRNA-Asn-GTT-9-2_R | TCTCTGTGGCGCAATCGGTT | | |
| tRNA-Gln-CTG-3-1_3-2_F | GTTCCATGGTGTAATGGTG | qRT-PCR | This study |
| tRNA-Gln-CTG-3-1_3-2_R | CGAGACTCGAACTCGGAT | | |
| tRNA-Gln-TTG-2-1_4-1_F | GGTCCCATGGTGTAATGGTT | qRT-PCR | This study |
| tRNA-Gln-TTG-2-1_4-1_R | CGAGATTCGAACTCGGAT | | |
| tRNA_Lys_TTT_2-1_F | CCTGGATAGCTCAGTCGGTAGAG | qRT-PCR | [41] |
| tRNA_Lys_TTT_2-1_R | GACTTGAACCCTGGACCCTCA | | |
| tRNA_Leu_CAA_1_2_F | CTCAAGCTTGGCTTCCTCGT | qRT-PCR | [41] |
| tRNA_Leu_CAA_1_2_R | GAACCCACGCCTCCATTG | | |
| PSMB6_F | CAAGAAGGAGGGCAGGTGTACT | qRT-PCR | [43] |
| PSMB6_R | CCTCCAATGGCAAAGGACTG | | |
| SDHA_F | CTGTCTTCATACGCTTCTGCACTC | qRT-PCR | [43] |
| SDHA_R | CCAGCCACTAGGTGCCAATC | | |
| COPS7A_F | CTGGCCACACTCATCCATC | qRT-PCR | [43] |
| COPS7A_R | AGGTAGAGGCAAAGTCACTCTCA | | |
| SS-70 | UCUAGUACUAUGUAUAUGAUAUCACACUCAGCACUACGUACUCAUAGC UAUGCUACACUAUGCACGCGAU | spike-in RNA for RNAseq | [44] |
| SS-94 | CGAAAUUAAUACGACUCACUAUAGGGGAAUUGUGAGCGGAUAACUGAC UGACUGACUAAAUAAUUUUGUUUAACUUUAAGAAGGAGAUAUACCA | spike-in RNA for RNAseq | [45] |

Supplementary Table 2: Detailed clinical and genetic characteristics of individuals with POLR9A pathogenic variants presented in the study.

| Subject | A-I.1 | A-II.1 | A-II.2 | B-II.1 | C-II.1 | D-II.1 | E-II.1 | F-I.1 | F-II.1 | G-II.7 | H-II.1 |
|---|---|---|---|---|---|---|---|---|---|---|---|
| **Identification** | | | | | | | | | | | |
| POLR9A variant | c.1078A>G p.(Arg360Gly) | c.1078A>G p.(Arg360Gly) | c.1078A>G p.(Arg360Gly) | c.2270C>T p.(Ser757Phe) | c.2278C>T p.(Arg760Cys) | c.2278C>T p.(Arg760Cys) | c.2357A>G p.(Lys786Arg) | c.2548C>A p.(His850Asn) | c.2548C>A p.(His850Asn) | c.2558C>T p.(Ala853Val) | c.2560G>A p.(Gly854Ser) |
| **Age at last examination** | | | | | | | | | | | |
| | 70–80 y | 40–50 y | 40–50 y | 30–40 y | 1–10 y | 11–20 y | 1–10 y | 51–60 y | 11–20 y | 11–20 y | 21–30 y |
| **Anthropometric data** | | | | | | | | | | | |
| Current Height (centile) | 178cm (between 50-75$^{th}$ centile) | 172cm (25$^{th}$ centile) | 188cm (between 91$^{st}$-98$^{th}$ centile) | 170cm (50$^{th}$ centile) | 145cm (between 3$^{rd}$-15$^{th}$ centile) | 165cm (between 3$^{rd}$-15$^{th}$ centile) | 122cm (50$^{th}$ centile) | 162.5 cm (3$^{rd}$ centile) | 160 cm (<3$^{rd}$ centile) | 157cm (20$^{th}$ centile) | n.d. |
| Current Weight (centile) | 70kg (between 50-75$^{th}$ centile) | 80kg (75$^{th}$ centile) | 105kg (>99.6$^{th}$ centile) | 55kg (50$^{th}$ centile) | 38kg (15$^{th}$ centile) | 37.5kg (<1$^{st}$ centile) | 25.4kg (between 75-85$^{th}$ centile) | 75.5 kg (between 75-90$^{th}$ centile) | 81.6 kg (>90$^{th}$ centile) | 35kg (<1$^{st}$ centile) | 36kg (<1$^{st}$ centile) |
| BMI | 22.1 | 27 | 29.7 | 19 | 18.1 | 13.78 | n.d. | 28.6 | 31.9 | 14.2 | n.d. |
| **Age at onset and first symptoms** | | | | | | | | | | | |
| Age at onset | 0–5 y | 0–5y | 11–15 y | 0–5 y | 0–5 y | 0–5 y | 0–5 y | 0–5 y | 0–5 y | 0–5 y | 0–5 y |
| First signs/symptoms | dragged feet when walked and often tripped | walking on tip toes | difficulty with walking, foot drop | unable to climb | could not sit upright | delayed walking | epilepsy | difficulty with walking and balance, falls | delayed walking, abnormal gait, poor balance, and falls | delayed walking | delayed walking, unsteady balance, falls |
| **Development** | | | | | | | | | | | |
| Global developmental delay | no | no | no | no | no | yes | yes | no | no | no | no |
| Regression | no | no | no | no | no | no | episodes of regression | no | no | no | no |
| Intellectual disability | no | no | no | no | no | yes | yes | no | no | no | no |
| Gross Motor | difficulty going up steps | persistent toe walking during childhood up until Achilles tendon operations during adolescence | normal | did not crawl, delayed walking, abnormal gait | delayed sitting, delayed walking | delayed walking, abnormal gait | delayed walking | age of walking uncertain, walking decline | delayed walking, abnormal gait | delayed walking | delayed sitting, delayed walking, abnormal gait |

| Subject | A-I.1 | A-II.1 | A-II.2 | B-II.1 | C-II.1 | D-II.1 | E-II.1 | F-I.1 | F-II.1 | G-II.7 | H-II.1 |
|---|---|---|---|---|---|---|---|---|---|---|---|
| Fine Motor | normal | normal | normal | normal at start, later function decreased | writes normally, can hold a pen / does not have the strength to open lids / has difficulty carrying objects | n.d. | no writing ability just raw drawings | normal | normal | normal | impaired since from an early age |
| Speech/ language development | normal | speech therapy needed for pronunciation | normal | normal | normal | normal | delayed speech | normal | normal | normal | normal |
| Neurological examination | | | | | | | | | | | |
| Central Nervous System | | | | | | | | | | | |
| Pyramidal signs and symptoms | no | no | no | no | n.d. | no | brisk tendon reflexes | no | no | no | no |
| Cerebellar ataxia | no | no | no | no | yes | no | no | no | no | no | yes |
| Other extrapyramidal signs | no | no | no | no | n.d. | no | no | no | no | no | no |
| Gait | walks independently - unstable | walks independently - unstable | walks independently - unstable | wheelchair - can help with transfers (11–20 y) | walks independently - unstable | wheelchair - can help with transfers | walks independently - unstable | wheelchair - can help with transfers | walks with assistance | walks independently - unstable | wheelchair - cannot help with transfers (11–20 y) |
| Tremor | n.d. | n.d. | n.d. | in hands around (0–10 y) | yes, in hands | no | no | no | no | no | no |
| Dysarthria | no | no | no | no | yes nasal speech | no | yes | yes | no | no | yes |
| Dysphagia | no | no | no | no | frequent choking - no longer chokes but everything must be cut very small / chews a lot and doesn't swallow | no | no | yes | no | no | yes |
| Epilepsy | no | no | no | no | no | no | yes, myoclonic and myoclonic-astatic epilepsy (onset during infancy) | no | no | no | no |

| Subject | A-I.1 | A-II.1 | A-II.2 | B-II.1 | C-II.1 | D-II.1 | E-II.1 | F-I.1 | F-II.1 | G-II.7 | H-II.1 |
|---|---|---|---|---|---|---|---|---|---|---|---|
| Brain MRI | n.d. | n.d. | n.d. | n.d. | normal (0–10 y) | n.d. | normal (0–10 y) | suggestive of chronic small vessel ischemic gliosis (51–60 y) | n.d. | n.d. | cerebellar atrophy (0–10 y) |
| Peripheral nervous system | | | | | | | | | | | |
| Muscle weakness – LL (MRC grade) | distal weakness hip flex 3/3; hip ext 3/3; hip abd 3/3; hip addu 3/3; knee flex 4/4; knee ext 4/4, ankle dorsiflex 0/0; plantar flex 0/0; plantar inv 0/0; plantar ever 0/0; great toe flex 0/0 great toe ext 0/0 | distal weakness ankle dorsiflex 0/0; plantar flex 0/0 | distal weakness ankle dorsiflex 0/0; | distal weakness 21–30 y: proximal muscles 2-3/2-3; ankle 0/0 | n.d. | distal weakness hip ext 5/5; hip flex 4/3; knee ext 3/3; knee flex 3/3; ankle dorsiflex 0/0; plantar flex 0/0 | distal weakness 1/1 | distal weakness 41–50 y: hip flexion 4/4; knee ext 4/4; ankle dorsiflex 0/0; plantar flex 0/0 at age 60 y: hip flexion 4/4; knee ext 3/3; ankle dorsiflex 0/0; plantar flex 0/0 | distal weakness 0–10 y: hip flex 5/5; knee ext 5/5; knee flex 5/5; ankle dorsiflex 0/0; ankle plantarflex 3/3; at age 20: hip flex 4/4; knee ext 4/4; knee flex 4/4; ankle dorsiflex 0/0; plantar flex 0/0 | distal weakness hips 5/5; knees 5/5; ankles 1/1; | distal weakness hips 3/3; knees 3/3; ankles 0/0 |
| Muscle weakness – UL (MRC grade) | distal weakness finger abd 4/4; | distal weakness finger abd 5/5; thumb abd 4/4; | distal weakness finger abd 4/4; thumb abd 4/4; | distal weakness 21–30 y: deltoideus 5/5; biceps 4/5; triceps 5/5; wrist ext 2/3; wrist flex 3/3; finger ext 0/0; finger abd 0/0 | n.d. | distal weakness shoulder abd 5/5; elbow flex 5/5; elbow ext 5/5; wrist ext 0/0; wrist flex 3/3; finger ext 0/0; finger flex 4/4; finger abd 0/0; APB 0/0 | 5/5 | distal weakness 41–50 y: shoulder abd 4/4; elbow flex 4/4; elbow ext 4/4; wrist ext 3/3, wrist flex 0/2; finger ext 0/0; finger abd 0/0; APB 2/2 51–60 y: shoulder abd 4/4; elbow flex 4/4; wrist ext 3/3, finger ext 0/0; finger abd 0/0 | distal weakness 0–10 y: shoulder abduct 5/5; elbow flex 5/5; elbow ext 4/4; wrist ext 5/5; wrist flex 4/4; finger ext 4/4; finger flex 4/4; finger abd 4/4; thumb abd 4/4; 11–20 y: shoulder abd 5/5; elbow flex 4/4; elbow ext 4/4; wrist ext 4/4; finger abd 0/0; thumb abd 0/0 | distal weakness; shoulders 5/5; elbows 5/5; wrist 5/5; intrinsic hand muscles 3-4/3-4 | distal weakness elbows 3/3; wrist 2/2; intrinsic hand muscles 0/0 |

| Subject | A-I.1 | A-II.1 | A-II.2 | B-II.1 | C-II.1 | D-II.1 | E-II.1 | F-I.1 | F-II.1 | G-II.7 | H-II.1 |
|---|---|---|---|---|---|---|---|---|---|---|---|
| Sensory superficial | pinprick: UL - impaired to mid forearm, LL - impaired to mid-thigh | pinprick: UL - impaired to fingertips, LL - impaired to upper thigh; temperature: UL - elbows, LL - above knee | pinprick, temperature and touch sensation to just above the ankles. | 21–30 y: pinprick reduced in hands | n.d. | n.d. | n.d. | 41–50 y: pinprick reduced to ankles and reduced to wrists<br><br>51–60 y: pinprick reduced to below knees | 0–10 y: Temperature (cold sensation) reduced below knees, normal at hands.<br><br>11–20 y: pinprick reduced in the feet. | reduced 2-point discrimination in fingers | reduced to ankles and to wrists |
| Proprioception | impaired at finger and toe joints | UL normal, LL lost to ankle | sensory ataxia | n.d. | n.d. | n.d. | n.d. | 41–50 y: absent at toes, reduced at ankles, normal at fingers | 0–10 y: normal at fingers/toes | reduced in LL | absent at toes, reduced at ankles, reduced at fingers |
| Vibration | UL - impaired to wrist, LL - impaired to iliac crest | UL - normal, LL - to knee | reduction to above the knees | n.d. | n.d. | reduced in right elbow and absent elsewhere | n.d. | 41–50 y: vibration sensation: Rydell Seiffer: absent to knees where it is reduced, and absent to the elbows;<br><br>51–60 y: vibration absent up to and including the knees | 0–10 y: vibration present but reduced at great toe and index finger.<br><br>11–20 y: vibration briefly present but very reduced at toes and fingers | reduced in LL | absent at toes and ankles, reduced at knees; reduced at fingers |
| Deep tendon reflexes | diffusely absent | diffusely absent | diffusely absent | diffusely absent | absent in distal LL | diffusely absent | n.d. | diffusely absent | diffusely absent | absent in LL and distal UL, reduced reflexes in proximal UL | absent in LL and distal UL, reduced reflexes in proximal UL |
| Skeletal Deformities | pes.planus | pes.planus | pes.planus | claw toes | pes.planus | hammer toes, limitations on elbow, knee and ankle flexion | kyphosis | pes.planus, hammer toes. | pes.planus | bilateral cavo_ varus.feet requiring orthopaedic surgery | kyphoscoliosis, pectus carinatum |
| Neurophysiology | | | | | | | | | | | |
| Age of evaluation | 71–80 y | 41–50 y | 41–50 y | 0–10 y | 0–10 y | 21–30 y | 0–10 y | 41–50 y | 21–30 y | 0–10 y | 0–10 y |

| Subject | A-I.1 | A-II.1 | A-II.2 | B-II.1 | C-II.1 | D-II.1 | E-II.1 | F-I.1 | F-II.1 | G-II.7 | H-II.1 |
|---|---|---|---|---|---|---|---|---|---|---|---|
| Motor: CMAP/ MCV/ Max onset latency | | | | | | | | | | | |
| Median | 0.1/27.9/16.0 (elbow) | 4.3/40/10.2 (elbow) | 5.0/38.9/12.5 (elbow) | n.a | MCV reduced to 50% | absent | n.d. | n.d. | 4.2/31/5.1 | 5.5/25.8/5.68 | 0.22/ 9/ 11 |
| Ulnar | 0.3/35.7/13.0 (above elbow) | 2.4/39/12.1 (above elbow) | 5.6/45.0/11.4 (above elbow) | n.a | MCV reduced to 60-70% | absent | n.d. | absent | n.d. | 3.9/25.2/5.16 | 0.1/ 16.5/ 6.8 |
| Peroneal | absent | absent | absent | n.a. | n.a. | absent | 3.5/24/9.58 | n.d. | n.d. | absent | absent |
| Tibial | absent | absent | absent | n.a. | n.a. | absent | 6.6 /29.5/7,97 | n.d. | n.d. | absent | absent |
| Sensory: SNAP/ SCV/ Max peak latency | | | | | | | | | | | |
| Median | absent | absent | absent | n.a. | absent | absent | n.d. | absent | n.d. | absent | absent |
| Ulnar | absent | absent | absent | n.a. | absent | absent | n.d. | absent | n.d. | absent | absent |
| Sural | absent | absent | absent | n.a. | n.d. | absent | n.d. | absent | n.d. | absent | absent |
| Other findings | | | | | | | | | | | |
| | n.d | n.d. | n.d. | described as having a demyelinating sensory-motor pattern | described as having a demyelinating sensory-motor pattern with secondary axonal degeneration | n.d. | n.d. | absent lateral antebrachial cutaneous SNAP; blink reflexes 19.2 msec | n.d. | n.d. | n.d. |
| Other neurological studies | | | | | | | | | | | |
| | n.d. | n.d. | n.d. | n.d. | n.d. | normal lumbar MRI; muscle biopsy with primary neurogenic abnormalities | EEG with epileptic activity. | n.d. | moderate obstructive sleep apnoea | n.d. | BAEP impairment at a high brainstem level |
| Biallelic POLR3-related phenotypes | | | | | | | | | | | |
| | no | no | no | teeth abnormalities: no enamel on teeth | GH deficiency: delayed bone age and shot stature responsive to GH treatment | teeth abnormalities: abnormally placed and shaped teeth | teeth abnormalities: tooth 74 is ankylosed and tooth 55 has delayed eruption | short stature | short stature | no | no |

| Subject | A-I.1 | A-II.1 | A-II.2 | B-II.1 | C-II.1 | D-II.1 | E-II.1 | F-I.1 | F-II.1 | G-II.7 | H-II.1 |
|---|---|---|---|---|---|---|---|---|---|---|---|
| Other (i.e. dysmorphisms, abnormalities in other systems) | | | | | | | | | | | |
| | excess atrial ectopic beats, mild gradual hearing loss | no | no | no | no | no | no | no | no | no | no |

Abbreviations are as follows: abd, abduction; addu, adduction; APB, Abductor pollicis brevis; BAEP, brainstem auditorial evoked potential; BMI, body mass index; CMTNS, Charcot-Marie-Tooth Neuropathy Score; CMAP, compound muscle action potential; EEG, electroencephalography; ext, extension; flex, flexion; GH, growth hormone; IQ, intelligence quotient; inv, inversion; LL, lower limbs; MCV, motor conduction velocity; MRC, Medical Research Council Scale for Muscle Strength; MRI, magnetic resonance imaging; mo, month or months; n.a., not available; n.d., not determined; SNAP, Sensory nerve action potential; SCV, sensory conduction velocity; UL, upper limbs; y, year or years. CMAP measured in millivolts (mV); MCV and SCV measured in meters per second (m/s); SNAP measured in microvolts (µV); Max peak latency measured in milliseconds (ms). Whenever available, height and weight charts from national health programs matching ancestry were used to determine the centiles. For other cases CDC growth charts were used as reference.

Supplementary Table 3: Summary of intra- and intermolecular interactions of POLR9A peripheral neuropathy-associated residues

| POLR3A residue | Intramolecular interactions | Intermolecular protein interactions | RSA % (Pol III assembly) | RSA % (Pol III assembly with nucleic acids) | Nucleic acid interaction |
|---|---|---|---|---|---|
| Arg360 | None | Gln1034 of POLR3B (PDB:7FJJ) - weak H-bond<br>Pro1035 of POLR3B (PDB: 7AE3) - weak vdW interaction<br>Thr1036 of POLR3B (PDB: 7AE3) - vdW interaction<br>Glu1037 of POLR3B (PDB: 7AE3) - vdW interaction<br>Gly1044 of POLR3B (PDB: 7AE3) - weak H-bond<br>Arg1046 of POLR3B (PDB: 7AE3) - vdW interaction<br>Ser1072 of POLR3B (PDB: 7AE3) - vdW interaction | 27.8 | 18.3 | Yes<br>dC23 of DNA (PDB: 7AE3) - H-bond |
| Ser757 | Leu753 - H-bond<br>Lys754 - weak H-bond<br>Arg760 - weak H-bond<br>Asp761 - H-bond | Asp63 of POLR3K (PDB: 8IUH) - vdW interaction | 22.6 | 22.6 | No |
| Arg760 | Leu756 - weak H-bond<br>Ser757 - weak H-bond<br>Ala763 - vdW interaction<br>Gly764 - H-bond | None | 19.5 | 19.5 | No |
| Lys786 | Gln543 - weak vdW interaction<br>Met780 - weak H-bond<br>Ala781 - weak H-bond | His932 of POLR3B (PDB: 7FJJ) - vdW interaction<br>Ser936 of POLR3B (PDB: 7FJJ) - vdW interaction<br>Glu89 of POLR3K (PDB: 7D59) - weak H-bond | 22.1 | 22.1 | No |
| His850 | Gln802 - H-bond<br>Phe838 - vdW interaction<br>Glu846 - H-bond<br>Phe847 - vdW interaction<br>Ala853 - weak H-bond<br>Gly854 - H-bond | Gln679 of POLR3B (PDB: 7AE3) - weak vdW interaction<br>Ser680 of POLR3B (PDB: 7AE3) - vdW interaction | 4.7 | 4.7 | No |
| Ala853 | Gln802 - weak vdW interaction<br>Phe849 - H-bond<br>His850 - weak H-bond<br>Glu856 - weak H-bond<br>Gly857 - weak H-bond | None | 7.4 | 7.4 | No |
| Gly854 | His850 - H-bond<br>Thr851 - vdW interaction<br>Gly857 - weak H-bond<br>Leu858 - H-bond | Ser86 of POLR3K (PDB: 7AE3) - weak H-bond | 2.4 | 2.4 | No |

Abbreviations are as follows: H-bond: hydrogen bond; vdW: van der Waals interaction. Interactions with DNA and interactions with residues in which missense variants are associated with peripheral neuropathy are highlighted in red.

Supplementary Table 4: Comparison of cardinal clinical features of POLR3-related monogenic disorders.

| | Monoallelic disorders | | Biallelic disorders | | |
|---|---|---|---|---|---|
| Gene | POLR9A | POLR9B | POLR9A | POLR9A?POLR9B? POLR7C?POLR9K? POLR9GL | POLR9A?POLR9B |
| Variant type | missense | missense | Non-canonical splicing or 5’UTR/ missense, PTV | missense, PTV | missense, PTV, non-canonical splicing site |
| Clinical diagnosis | Demyelinating PN with variable CNS and non-neurological involvement | CMT 1I, epileptic encephalopathy | Cerebellar ataxia and/or spastic paraplegia | POLR3-Related Leukodystrophy | Wiedemann-Rautenstrauch syndrome |
| Total number of individuals | 11 | 23 | 48 | 105 | 18 |
| Sensorimotor peripheral neuropathy[a] | 100% (11/11) | 81% (13/16) | 1/30[c] | n.d.[d] | n.d. |
| Global developmental delay / Intellectual disability | 18.2% (2/11) | 77.3% (17/22) | 4.2% (2/48) | 52.4% (54/103) | 70% (7/10) |
| Epilepsy | 9.1% (1/11) | 68.2% (15/22) | 2.1% (1/48) | 19.2% (19/99) | 12.5% (2/16) |
| myoclonic epilepsy | 9.1% (1/11) | 50% (11/22) | n.d. | n.d. | n.d. |
| Cerebellar signs | 18.2% (2/11) | 40.9% (9/22) | 91.7% (44/48) | 99% (104/105) | 25% (4/16) |
| Cerebellar atrophy | 9.1% (1/11) | 14.3% (3/21) | 30.8% (12/39) | 90.7% (88/97) | 0/7 |
| Pyramidal signs | 10% (1/10) | 45.5% (10/22) | 75% (36/48) | common[e] | 56.2% (9/16) |
| Other extrapyramidal signs | 0/11 | 0/22 | 25% (12/48) | uncommon[f] | 0/18 |
| Hypomyelinating leukodystrophy | 0/4 | 0/22 | 0/35 | 95.9% (93/97) | 28.6% (2/7) |
| Microcephaly | 0/1 | 44.4% (8/18) | n.d. | n.d. | 66.6% (10/15) |
| Low weight | 30% (3/10) | 5.5% (1/18) | 7.7% (1/13) | n.d. | 100% (9/9) |
| Short stature | 30% (3/10) | 11.1% (2/18) | 5.3% (1/19) | 51.6% (47/91) | 85.7% (6/7) |
| Endocrine abnormalities | 9.1% (1/11) | 13% (3/23) | 2.1% (1/48) | 75.8% (47/62) | 44.4% (4/9) |
| Dental abnormalities | 27% (3/11) | 0/22 | 68.4% (26/38) | 87.1% (88/101) | 50% (8/16) |
| Lipodystrophy | 0/11 | 0/22 | 0/48 | 0/105 | 100% (18/18) |
| WRS facial phenotype[b] | 0/11 | 0/22 | 0/48 | 0/105 | 100% (18/18) |
| Data source | This study | [46-53] | [54-56] | [57,58] | [59] |

Abbreviations are as follows: 5’UTR, 5’ upstream untranslated region; n.d. – not determined; NCS- nerve conduction studies; PTV – protein truncating variant; WRS – Wiedemann-Rautenstrauch syndrome.

[a] only individuals with NCS reported were considered.

[b] WRS facial phenotype is characterized by sparse hair, triangular face, thin upper vermilion, small mouth, pointed chin.

[c] reported in an individual with a homozygous hypomorphic variant.

[d] Gauquelin et al. reports that individuals with POLR3-related disorders have normal NCS.

[e] number of affected individuals in the cohort is not specified, but pyramidal signs were described as usually absent in young children and developed slowly in older patients.

[f] number of affected individuals in the cohort is not specified, but the authors report that few patients present extrapyramidal signs, mainly dystonia.

Supplementary Table 5: Interpretation of peripheral neuropathy-associated variants according to American College of Medical Genetics and Genomics recommendations.

| Variant | ACMG criteria | ACMG classification |
|---|---|---|
| c.1078A>G, p.Arg360Gly | PP2, PP3(moderate), PM2, PS3 | Likely pathogenic |
| c.2270C>T, p.Ser757Phe | PP2, PP3(moderate), PM2, PS2, PS3 | Pathogenic |
| c.2278C>T, p.Arg760Cys | PP2, PP3(moderate), PM2, PS2 | Likely Pathogenic |
| c.2357A>G, p.Lys786Arg | PP2, PP3, PM2, PS2, PS3 | Pathogenic |
| c.2548C>A, p.His850Asn | PP2, PP3(moderate), PM2, PS2, PS3 | Pathogenic |
| c.2558C>T, p.Ala853Val | PP2, PM2, PS2, PS3 | Pathogenic |
| c.2560G>A, p.Gly854Ser | PP2, PM2, PS2, PS3 | Pathogenic |
